# Parkinson’s disease-associated genetic variants synergistically shape brain networks

**DOI:** 10.1101/2022.12.25.22283938

**Authors:** Zhichun Chen, Bin Wu, Guanglu Li, Liche Zhou, Lina Zhang, Jun Liu

**Affiliations:** Department of Neurology and Institute of Neurology, Ruijin Hospital affiliated to Shanghai Jiao Tong University School of Medicine, Shanghai, 200025, China; Department of Neurology, The Second Xiangya Hospital, Central South University, 139 Renminzhong Road, Changsha, 410011, China; Department of Neurology, Xuchang Central Hospital affiliated with Henan University of Science and Technology, Henan, 461000, China; Department of Biostatistics, Shanghai Jiao Tong University School of Medicine, Shanghai, 200025, China

**Keywords:** Parkinson’s disease, genetic variant, brain networks, graphical analysis, clinical manifestations.

## Abstract

**Background:** Over 90 genetic variants have been found to be associated with Parkinson’s disease (PD) in genome-wide association studies, however, the neural mechanisms of previously identified risk variants in PD were largely unexplored. The objective of this study was to evaluate the associations between PD-associated genetic variants and brain gene expressions, clinical features, and brain networks.

**Methods:** PD patients (n = 198) receiving magnetic resonance imaging examinations from Parkinson’s Progression Markers Initiative (PPMI) database were included in the analysis. The effects of PD-associated genetic variants assayed in PPMI database on clinical manifestations and brain networks of PD patients were systematically evaluated.

**Findings:** Most associations between 14 PD-associated risk variants and clinical manifestations of PD patients failed to reach the stringent *p*-value threshold of 0.00026 (0.05/14 clinical variables x 14 variants). Shared and distinct brain network metrics were significantly shaped by PD-associated genetic variants. Small-worldness properties at the global level and nodal metrics in caudate and putamen of basal ganglia network were preferentially modified. Small-worldness properties in gray matter covariance network mediated the effects of *OGFOD2/CCDC62* rs11060180 on motor assessments of PD patients.

**Interpretation:** Our findings support that both shared and distinct brain network metrics are shaped by PD-associated risk variants. Small-worldness properties modified by *OGFOD2/CCDC62* rs11060180 in gray matter covariance network are associated with motor severity of PD patients. Future studies are encouraged to explore the underlying mechanisms of PD-associated risk variants in PD pathogenesis.

**Funding:** This work was supported by grants from the National Key Research and Development Program (2016YFC1306505) and the National Natural Science Foundation of China (81471287, 81071024, 81171202).

## Introduction

Parkinson’s disease (PD) is the most common movement disorder and the second most common neurodegenerative disease in the world. Since PD was first reported in 1817, the study of PD has a history of more than 200 years. However, PD patients remain incurable now due to the limited understanding of PD pathogenesis. Recent studies have shown that genetic variation is a fundamental mechanism to drive the initiation and progression of PD.^1–9^ Additionally, a rapidly growing number of genes associated with PD risk have been detected, including *GPNMB* and *MAPT*.^10–12^ However, the specific roles of previously identified risk genes in PD pathogenesis are still unclear. With the discovery of new risk genes in PD, in recent years, deciphering the biological functions of PD-associated risk genes and their roles in the occurrence and progression of PD at the molecular, cellular, neural circuit, and behavior levels has continuously driven the new research breakthroughs in PD.^2–9^ Over the past decade, both the physiological and pathological functions of a range of PD-associated risk genes have been dissected, including *SNCA*,^7,13–15^ *Parkin*,^2–5^ *PINK1*,^4–6^ *LRRK2*,^16–21^ *TMEM175*,^9^ and *ATP13A2*.^8^ These breakthroughs deepen our understanding of the neurodegenerative process of PD and lay the foundation for the exploration of new therapeutic targets.

Currently, one of our main challenges is to investigate the biological functions and pathogenic or protective roles of numerous single nucleotide polymorphisms (SNPs) that have been identified in PD. However, it is time-consuming and laborious to study the biological functions and its specific roles in disease pathogenesis for each PD-associated risk SNP. Previously, several SNPs have been reported to modify clinical features of PD patients, including motor symptoms, non-motor symptoms, striatal dopamine transporter (DAT) uptake, and biomarkers in cerebrospinal fluid (CSF).^11,20, 22–29^ Through expression quantitative trait loci (eQTL) analysis, studies have also reported some SNPs conferring risk of PD were associated with brain gene expressions.^10,30–32^ Due to these findings, there is no doubt that those SNPs significantly associated with clinical features and brain gene expressions are more likely to be functional genetic variants in PD. Therefore, with the eQTL analysis and association analysis between clinical assessments and genotypes of PD-associated risk SNPs, we can screen out more meaningful SNPs that deserved to be further investigated.

Over the past three decades, the methodologies of functional magnetic resonance imaging (fMRI) have been utilized to identify the structural or functional presentations associated with specific genetic variants.^33–35^ In addition, recent studies have combined the functional and structural network analysis to identify network alterations associated with genetic variants and clinical features of diseases.^36–39^ In familial PD, PD-associated mutated genes, such as *GBA* and *LRRK2*, have been shown to significantly modify the brain networks.^40–42^ Recently, we showed that a PD-associated risk SNP, *MAPT* rs17649553, is associated with small-world topology of structural network in PD.^43^ Nevertheless, whether PD-associated risk genes affected brain networks in sporadic PD remain largely elusive. Accumulated evidence has shown that network metrics in both functional network and structural network were significantly associated with motor and non-motor symptoms of PD patients.^44–48^ Therefore, brain networks modified by PD-associated risk genes may contribute to the heterogeneity of clinical features of PD patients.^43^

In this study, our hypothesis is that brain network metrics can be shaped by PD-associated risk genes and mediate the effects of PD-associated genetic variants on clinical manifestations of PD patients. We determine to combine the eQTL analysis, clinical phenotype association analysis, and neuroimaging analysis to respectively assess how the PD-associated risk genes affect the brain gene expressions, clinical manifestations, and brain network metrics in PD patients from Parkinson’s Progression Markers Initiative (PPMI) database (www.ppmi-info.org/data). Specifically, our objectives include: (i) to identify PD risk-associated SNPs with significant eQTL effects from 72 SNPs assayed in PPMI database. (ii) to examine whether PD-associated risk variants were associated with the clinical assessments of PD patients; (iii) to determine whether PD-associated genetic variants modify the brain functional and structural network metrics; (iv) to explore the associations between brain network metrics and clinical assessments of PD patients; (v) to investigate whether the functional and structural network metrics mediate the effects of PD-associated genetic variants on clinical assessments of PD patients.

## Methods

### Participants

The data used in the preparation of this Article were obtained from PPMI database, which is a robust open-access online resource platform, providing clinical, imaging, ‘omics, genetic, sensor, and biomarker data set to deepen the understanding of PD and speed scientific breakthroughs and new treatments. The data collection protocols for PPMI have been published and available online (http://www.ppmi-info.org/).^49,50^ The PPMI study was approved by institutional review boards at each participating site and was conducted in accordance with the Declaration of Helsinki and the Good Clinical Practice (GCP) guidelines after approval of the local ethics committees of the participating sites. All participants completed written informed consent before they were enrolled into PPMI study. The inclusion criteria for participants in PPMI cohort has been published previously.^50^ Specifically, PD participants were included if they met the criteria below: (i) The participant was diagnosed with PD based on the diagnostic criteria of International Parkinson and Movement Disorder Society;^51^ (ii) The participant underwent 3D T1-weighted MPRAGE imaging or resting-state fMRI or diffusion tensor imaging (DTI); (iii) The participants received whole exome sequencing on DNA samples extracted from the whole blood; (iv) The participants didn’t carry genetic mutations of familial PD demonstrated by whole exome sequencing; (v) The participants were not diagnosed with other neurological diseases except PD; (vi) The participants had no evidence of structural abnormalities in T1-weighted or T2-weighted images, which were visually inspected by the investigators of this study (Jun Liu and Zhichun Chen). The PD patients were excluded if they were diagnosed with dementia or treated with neuroleptics, anticoagulants, metoclopramide, α-methyldopa, methylphenidate, reserpine, or amphetamine derivative. Finally, a total of 198 PD patients were identified to have 3D T1 images. Of these PD patients, one hundred and forty-six of them underwent DTI examinations and 83 patients having DTI images also received resting-state fMRI. In addition, almost all the patients performed iodine-123-labelled ioflupane SPECT to measure DAT activity. Volume of interest were placed on the left and right caudate, left and right putamen, and the occipital cortex (reference tissue) to extract the count densities for each of the four striatal regions.^50^ Striatal binding ratios (SBR) of caudate and putamen were calculated as (target region/reference region)-1.^50^ We downloaded all the SBR data from the PPMI database. Most of the PD patients also got tested for indices in CSF, including levels of β-amyloid (Aβ), α-syn (α-synuclein), p-tau, and tau, which were also downloaded from PPMI database. The control participants were included if they met the inclusion criteria below: (i) They were 30 years old or above; (ii) They received clinical assessments that PD participants examined; (iii) They were relatively healthy and not diagnosed with active, clinically manifested neurological disease. The exclusion criteria for control participants include: (i) They had first-degree relatives with PD; (ii) They carried genetic mutations of PD; (iii) They were treated with neuroleptics, anticoagulants, metoclopramide, α-methyldopa, methylphenidate, reserpine, or amphetamine derivative. Both PD and control participants were not from the genetic PPMI cohort and prodromal cohort. Because only a small percentage of control participants received MRI examinations, they were not included in the neuroimaging analysis. To ensure consistency in the timing of data collection, we kept the maximum time gap for all types of data collection within 3 months. The basal demographical data, motor symptoms, non-motor symptoms, SBR, and CSF indices were collected for 198 PD patients and 189 control participants (Table 1). Unpaired *t* test or *X*^2^ test was used for the comparison of clinical variables between control and PD patients. The demographical and clinical data were not statistically different among those with 3D T1 images, or DTI images, or resting-state images (Table S1). One-way ANOVA test was used for the comparisons of clinical variables among three groups. To investigate how PD-associated risk genes affect clinical assessments and brain networks, the genotypes of 72 SNPs examined by whole-genome sequencing in PPMI database were downloaded and investigated (http://www.ppmi-info.org/). The study flowchart of key risk variants from 72 SNPs was shown in Figure 1. Among 72 SNPs, fourteen SNPs both conferring risk for PD and exerting significant eQTL effects were specifically selected and their effects on clinical assessments and brain network metrics were systematically evaluated (Fig. 1).

**Figure 1.**
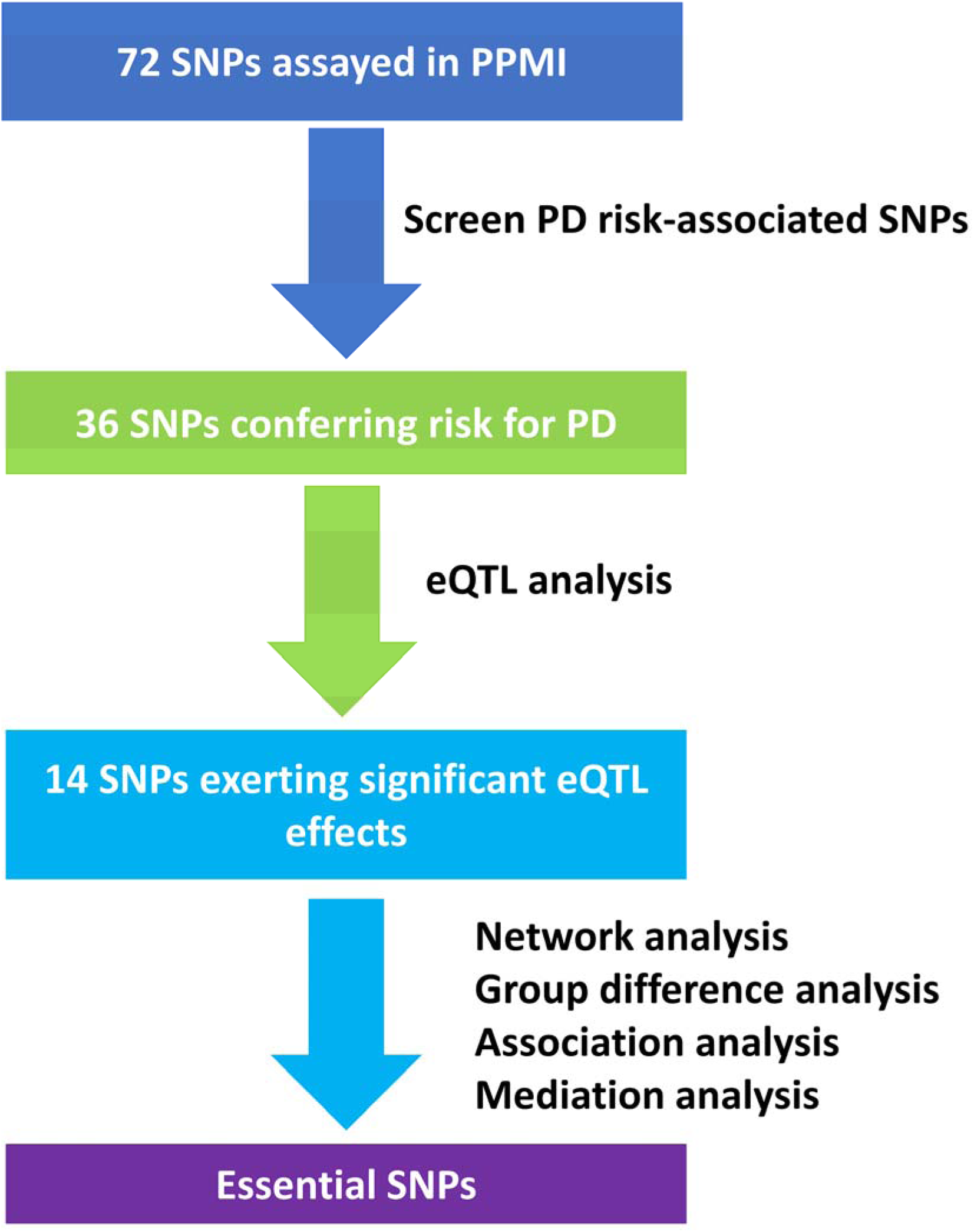
The study flowchart of key risk variants from 72 SNPs assayed in PPMI database.

### Image acquisition

All the fMRI images were acquired on 3T Siemens MRI scanners (either Trio™ or Verio™ systems, Siemens Healthcare, Malvern, PA) at different participating site. Most of 3D T1 MPRAGE sequence was scanned with following parameters: TR = 2300 ms, TE = 2.98 ms, Inversion time = 900 ms, Voxel size = 1 mm^3^, Matrix = 240 x 256 mm, Flip Angle = 9°, Slice thickness =1.2 mm. The resting-state fMRI was acquired for 8 minutes and 24 seconds with parameters of TR = 2400 ms, TE = 25 ms, Voxel size = 3.3 mm^3^, Field of View = 222 mm, Flip Angle = 80°, Slice thickness = 3.3 mm. Single shot echo-planar imaging (EPI) sequence was used for DTI with the following parameters ^52^: TR= 8,400-8,800 ms, TE = 88 ms, Voxel size = 2 mm^3^, twofold acceleration, Slice thickness = 2 mm. The DTI images were performed along 64 sensitization directions with a b-value of 1000 s/mm^2^. The MRI protocols were electronically distributed to each participating site to guarantee consistent installations. During the resting-state imaging, participants were required to stay quietly with clear mind and not to sleep.

### Image preprocessing

The processes of structural and functional network analysis were shown in Figure 2. Before all the images were preprocessed, the DICOM format images were transformed into NIFTI format images. For the preprocessing of resting-state fMRI images, the first 10 volumes were removed to achieve magnetization equilibrium. The remaining images were then slice-timing corrected and realigned. The rest of datasets were spatially normalized to Montreal Neurological Institute (MNI) space using EPI template in SPM12 (https://www.fil.ion.ucl.ac.uk/spm/software/spm12/). A 4-mm Gaussian smoothing kernel was applied to improve anatomical variance and signal to noise ratio. All voxel time series were then bandpass filtered (0.01∼0.1 HZ) to emphasize the low-frequency correlations and eliminate the low-frequency drift and high-frequency noises. To adjust the head motions, six head motion parameters, signals of white matter and CSF, were regressed out from each dataset. Among 83 PD participants with resting-state fMRI, we excluded 9 participants with head motions frame-wise displacement > 0.5 mm and head rotation > 2°. The remaining 74 PD participants underwent functional network analysis.

**Figure 2.**
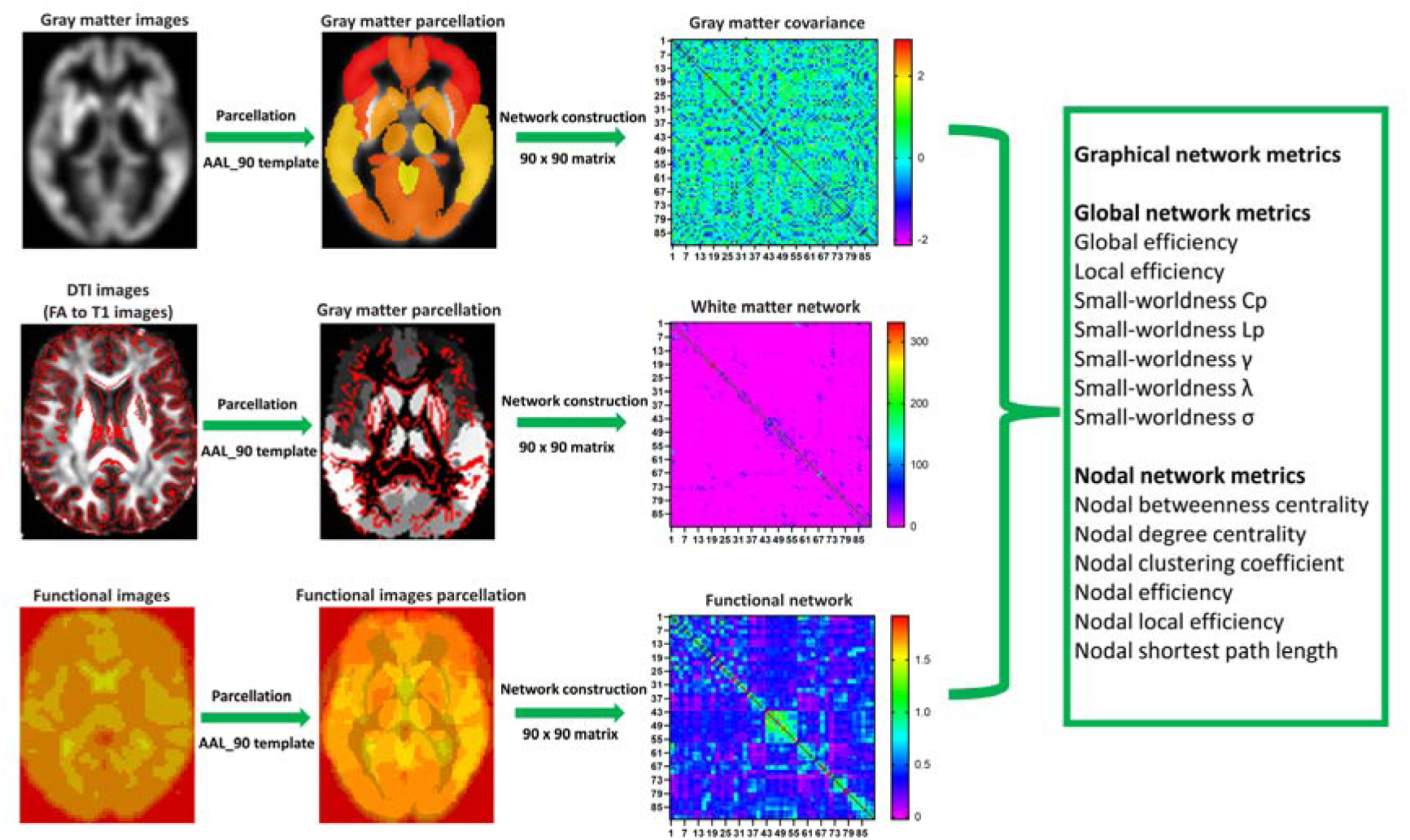
The flowchart of structural and functional network analysis in the present study. The 90 x 90 matrices of gray matter, white matter, and functional networks were derived from T1 images, DTI images, and resting-state functional images, respectively. The global network metrics and nodal network metrics were computed using GRETNA toolbox.

For DTI analysis, we included all 146 PD participants. Their DTI images were preprocessed using the FMRIB Software Library toolbox (FSL, https://fsl.fmrib.ox.ac.uk/fsl/fslwiki). The images preprocessing included the following major steps: (i) Removing the nonbrain tissue and extracting the brain using the Brain Extraction Tool; (ii) Correcting the eddy-current distortion and head motions using eddy_correct command; (iii) Computing the diffusion tensor metrics using dtifit command. (iv) Normalizing the images into MNI space.

The gray matter images of 198 PD participants were preprocessed using voxel-based morphometry (VBM) methodology ^53^ implemented on CAT12 toolbox ^54^ running within SPM12. Briefly, the T1 images were firstly normalized with Di[eomorphic Anatomic Registration Trough Exponentiated Lie algebra algorithm template and then segmented into gray matter, white matter, and CSF. The segmented gray matter images were further smoothed before the construction of gray matter covariance network. For surface-based morphometry (SBM), a fully automated method was used to measure cortical thickness and reconstruct the central surface using CAT12 toolbox. Partial volume information, sulcal blurring and asymmetries were handled by projection-based thickness. The processed surface data were smoothed using 15 mm kernel (12∼18mm kernels are widely used for SBM) before statistical analysis.

### Network construction

The widely-used Automated Anatomical Labelling (AAL) atlas with 90 regions provided reliable and unbiased localizations of regions of interest (ROIs) with a good anatomical interpretability,^55,56^ thus, AAL-90 template was used to define 90 cortical and subcortical nodes. The functional connectivity was calculated using the Pearson’s correlation of mean time courses of the 90 nodes. To improve the normality, the Fisher’s r-to-z transformation was performed. This created a 90 x 90 functional connectivity matrix for each subject.

Deterministic tractography was performed for individual preprocessed DTI images based on AAL atlas using the Fiber Assignment by Continuous Tracking (FACT) algorithm embedded in a free open Matlab toolbox PANDA (http://www.nitrc.org/projects/panda/). To construct white matter fibers, streamlines were created in each voxel along the principle diffusive direction. When the streamline reached a voxel with fractional anisotropy (FA) < 0.2 or tract angle > 45°, the streamline tracking was terminated. We used the fiber number (FN) to define structural connectivity. We didn’t set the threshold for FN within network here, because the threshold didn’t significantly affect the network properties. Finally, a 90 x 90 white matter FN matrix was constructed for each participant.

The methodology to construct the gray matter covariance network has been described previously.^57,58^ In brief, the AAL atlas was used to define the 90 subcortical and cortical nodes with 3 x 3 x 3 voxels. The network edges were computed as the correlation coefficients of gray matter volume between each pair of 90 nodes. The correlation coefficients were then Fisher’s r-to-z transformed. Finally, a 90 x 90 gray matter covariance matrix was created for individual participant.

### Network graphical analysis

The graph theory was used to analyze the topological properties of above three types of networks. We calculated both global and nodal metrics of networks using GRETNA toolbox (https://www.nitrc.org/projects/gretna/). The global network metrics included global efficiency, local efficiency, and small-worldness properties: clustering coefficient (Cp), characteristic path length (Lp), normalized clustering coefficient (γ), normalized characteristic path length (λ), and small worldness (σ). The nodal metrics estimated in this study included nodal betweenness centrality, nodal degree centrality, nodal efficiency, nodal local efficiency, nodal Cp, and nodal shortest path length. We calculated the area under curve (AUC) for each network metric. The definitions and computations of above topological parameters were described below and could also be found in previous studies.^59,60^ We defined *N* as the set of all nodes in a network, *n* as the number of nodes, *L* as the set of all edges (links) in the network, and *l* as the number of edges. (*i*, *j*) is a link between nodes *i* and *j* (*i*, *j* ∈ *N*). *a_ij_* is the connection status between node *i* and node *j*. *a_ij_* =1 when the link between node *i* and node *j* exists, *a_ij_* = 0 otherwise (*a_ij_* = 0 for all *i*). The number of links is computed as *l* = ∑*_i,j_*_∈*N*_ *a_ij_*.

The degree (*k_i_*) is defined as the number of links connected to a node *i*, then the degree of this node *i* is computed as below:

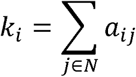

The shortest path length between node *i* and node *j* is computed as below:

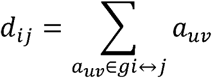

*gi* ↔ *j* is the shortest path between node *i* and node *j*.

The nodal shortest path length is the average shortest path length between node *i* and all other nodes. It is computed as below:

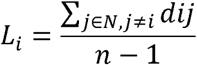

The node local efficiency measures the efficiency of local information transfer in node *i*. The node local efficiency is computed as below:

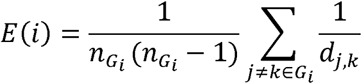

*G_i_* is the Graph composed of neighbor nodes of node *i*. *d_j,k_* represents shortest path length between node *j* and node *k*.

The node efficiency measures the efficiency of information transfer for node *i* in the network. Node efficiency is computed as below:

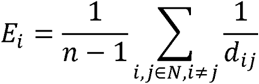

*d_i,j_* represents shortest path length between node *i* and node *j*.

The global efficiency is defined as the average *E_i_* of all nodes in a network. It is computed as below:

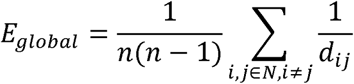

The local efficiency is defined as the average *E(i)* of all nodes in a network. It is computed as below:

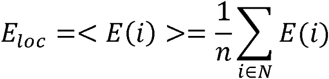

The node Cp measures how closely nodes in a network tend to group together. It is computed as below:

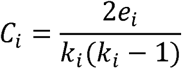

The network Cp is the average node Cp of a network and is computed as below:

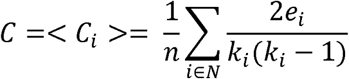

Characteristic path length (Lp) of the network is defined as the average shortest path length of all nodes in a network. It is computed as below:

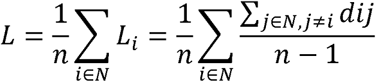

where *Li* is the average shortest path length between node *i* and all other nodes.

The small-worldness *γ* is computed as below:

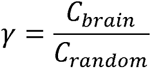

where *C_brain_* and *C_random_* represents the Cp of brain network and random network, respectively.

The small-worldness *λ* is computed as below:

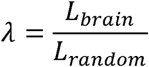

where *L_brain_* and *L_random_* represents the *L* (Characteristic path length) of brain network and random network, respectively.

The small-worldness *σ* is computed as below:

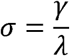

### The acquisition of eQTL data

All the eQTL data of analyzed SNPs were obtained from the Genotype-Tissue Expression (GTEx, https://www.gtexportal.org/home/) Project. The normalized effect size (NES) was computed as the effect of the alternate allele relative to the reference allele (https://www.gtexportal.org/home/faq). Uncorrected *p* < 0.05 was reported to identify the potential eQTL effect. A smaller *p* < 0.0001 was reported as higher eQTL effect.

### Statistical Analysis

#### Association analysis between clinical assessments and genotypes

The associations between genotypes of SNPs and clinical features of PD patients were analyzed using multivariate regression models. During the multivariate regression analysis, the dependent variables (n=14) included the scores of Unified Parkinson’s Disease Rating Scale part III (UPDRS-III), Epworth Sleepiness Scale (ESS), Geriatric Depression Scale, REM Sleep Behavior Disorder Screening Questionnaire (RBDSQ), Scale for Outcomes in Parkinson’s Disease-Autonomic (SCOPA-AUT), Benton Judgment of Line Orientation (BJLOT), Letter Number Sequencing, Montreal Cognitive Assessment (MoCA), Hopkins Verbal Learning Test-Revised (HVLT-R: Total Recall and Immediate Recall), Semantic Fluency Test, and SBRs (bilateral striatum), as well as α-syn level in CSF. The independent variables were the genotypes of 14 SNPs. During the multivariate regression analysis, age, sex, disease duration, and genotypes of remaining 13 SNPs were included as covariates and *β* (coefficients) and *p-v*alues were reported for each SNP. Because of the lack of total levodopa equivalent daily dose (LEDD) data in most PD patients, LEDD was not included as a covariate during the association analysis. The multivariate regression models were shown below (*SNP_1_*, *SNP_2_*, …, *SNP_i_* represent the genotypes of SNPs):

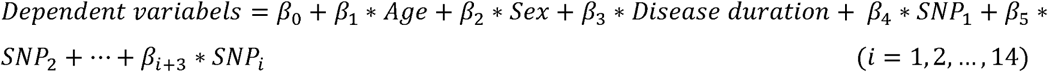

Results with *p <* 0.05 and *p <* 0.00026 (0.05/14*14; 14 clinical variables x 14 SNPs) after Bonferroni correction for multiple statistical tests were reported.

#### Comparisons of clinical assessments among different genotype configurations

Unpaired t-test (for two groups comparison) and one-way ANOVA test followed by Bonferroni post-hoc test (for three or more groups comparison) were used to compare the difference of clinical assessments among different SNP configurations. *p <* 0.05 was considered statistically significant.

#### Comparisons of graphical network metrics

The group differences of global graphical network metrics (global efficiency, local efficiency, and small-worldness properties) were analyzed using two-way ANOVA test followed by Bonferroni post-hoc test. Bonferroni-corrected *p* < 0.0036 (0.05/14; 14 SNPs) was considered statistically different for global network metrics. The nodal network metrics (nodal betweenness centrality, nodal degree centrality, nodal efficiency, nodal local efficiency, nodal Cp, and nodal shortest path length) of 90 nodes in AAL atlas were compared between 2 or among 3 genotype groups of 14 SNPs using two-way ANOVA test followed by Bonferroni post-hoc test. Uncorrected and Bonferroni-corrected results were reported based on the number of network metrics and SNPs.

#### Association analysis between clinical assessment or genotypes and network metrics

The univariate correlation analysis between scores of clinical assessments and graphic network metrics was conducted by Pearson correlation method. The multivariate regression models were also used for the association analysis between scores of clinical assessments or genotypes of risk SNPs and graphical network metrics. Before regression analysis, IBM SPSS Statistics Version 26 was used to check whether multicollinearity occurred and no multicollinearity was found. Age, sex, years of education, and disease duration were included as covariates during multivariate regression analysis. Results with uncorrected *p <* 0.05 and Bonferroni-corrected results were reported.

#### Comparisons of gray matter volume or cortical thickness

The whole brain comparison of gray matter volume or cortical thickness among different genotype groups was performed using ANOVA models in CAT12 toolbox and SPM12. The age, sex, and disease duration were included as covariates. *p <* 0.05 after Bonferroni correction was considered statistically different.

#### Comparisons of integrity of white matter tracts

The comparison of integrity (FA) of white matter tracts among different genotype groups was performed using ANOVA models. The age, sex, and disease duration were included as covariates. *p <* 0.05 after Bonferroni correction was considered statistically different.

#### Mediation analysis

IBM SPSS Statistics Version 26 was utilized to perform mediation analysis. The independent variable in the mediation model was genotypes of *OGFOD2/CCDC62* rs11060180, *GCH1* rs11158026, and *ZNF646/KAT8/BCKDK* rs14235. The dependent variable was UPDRS-III scores. The mediators were small-worldness γ and σ of gray matter covariance network. We modeled the mediated relationships (indirect path) between genotypes and UPDRS-III scores. The model also included the direct path from genotypes to the UPDRS-III scores. During the mediation analysis, age, sex, disease duration, and years of education were included as covariates. *p* < 0.05 was considered statistically significant.

#### Cross-validation analysis

K-fold cross-validation analysis was performed using R version 4.3.0 to evaluate the robustness of the regression models in this study. Statistical metrics derived from cross-validation analysis included root mean squared error (RMSE), coefficient of determination (R²), and mean absolute error (MAE). These metrics were used to assess the statistical robustness of the regression models.

## Results

### Demographical variables

The clinical data of 198 PD patients and 189 control participants were shown and compared in Table 1.

### The identification of 14 key risk SNPs in PD

The study flowchart of key risk variants from 72 SNPs assayed in PPMI database was shown in Figure 1. The selected key risk SNPs exhibited the characteristics shown below: (i) The SNP was associated with PD risk according to previous genetics studies;^10,11, 61–63^ (ii) The SNP showed significant eQTL effect according to GTEx database (https://www.gtexportal.org/home/).

Initially, we identified 36 SNPs conferring risk of PD from 72 SNPs examined in PPMI database according to previous literature, SNP database of NCBI (https://www.ncbi.nlm.nih.gov/snp/), and PDGene database (http://www.pdgene.org/). With the exception of *COMT* rs4680, which was shown to be associated with PD in small samples, all remaining 35 SNPs have been reported as risk variants of PD in previous large-scale genome-wide association studies.^10,11, 61–63^ For example, recent studies have confirmed that *GPNMB* rs199347 was significantly associated with PD.^10,12^

Based on the GTEx Project (https://www.gtexportal.org/home/), we checked whether 36 SNPs associated with PD risk were significantly correlated with gene expressions in multiple brain regions. Of 36 SNPs assessed, a total of 14 key risk SNPs were shown to be associated with brain gene expressions. The 14 key risk SNPs were located in 14 loci of the genome, including *OGFOD2/CCDC62* rs11060180, *GCH1* rs11158026, *ZNF646/KAT8/BCKDK* rs14235, *COMT* rs4680, *BIN3* rs2280104, *NUCKS1/Rab7L1* rs823118, *MAPT* rs17649553, *LRRK2* rs76904798, *GALC/GPR65* rs8005172, *ZNF184* rs9468199, *FAM47E/STBD1* rs6812193, *TMEM163* rs6430538, *NCKIPSD* rs12497850, and *GPNMB* rs199347 (Table 2 and Table S1). The genotype distribution of 14 key risk SNPs in our participants didn’t deviate Hardy-Weinberg equilibrium (all *p >* 0.05, *X*^2^ test). In addition, the distribution of genotype was not significantly different with regard to age, sex, and disease duration (*p >* 0.05). Of 14 key risk SNPs, six of them, including *MAPT* rs17649553, *ZNF646/KAT8/BCKDK* rs14235, and *NCKIPSD* rs12497850, dramatically modified the gene expressions of multiple brain regions, including caudate, hippocampus, substantia nigra, and cortex according to GTEx project (Fig. S1). To understand the biological pathways and cellular functions associated with 14 SNPs, we used STRING database to construct the protein-protein interaction (PPI) network of differentially expressed genes and performed functional enrichment analysis to reveal the biological processes, local network cluster, cellular components, and molecular functions associated with the PPI network. As shown in Figure S2, we found these SNPs were associated with molecular pathways involved in endocytosis, autophagy, lysosome, mitochondria metabolism, nuclear pore complex, gene transcription, and synaptic vesicles.

### Associations between 14 key risk SNPs and clinical variables

The associations between 14 key risk SNPs and clinical variables were performed using multivariate regression analysis. The results were shown in Table 2. We found most associations between genotypes and clinical variables had low effect sizes and did not reach the stringent Bonferroni-corrected *p* threshold of 0.00026 (0.05/14 clinical variables x 14 SNPs), except that *GCH1* rs11158026 was significantly associated with striatal binding ratios of bilateral striatum (β = -0.16, *p <* 0.00026).

### Group differences of clinical assessments among different genotypes of individual SNP

We have revealed some SNPs were significantly associated with scores of clinical assessments as shown above (Table 2), then we examined whether scores of clinical assessments were significantly different among different genotype groups. As shown in Figure S3, *OGFOD2/CCDC62* rs11060180 G-carriers showed higher UPDRS-III scores compared to AA carriers (*p* < 0.05; Fig. S3a). Consistently, UPDRS-III scores in GG carriers were higher than those of AA (*p* < 0.01) and AG carriers (*p* = 0.0744). Similarly, *GCH1* rs11158026 T-carriers exhibited higher UPDRS-III scores compared to CC carriers (*p* < 0.05; Fig. S3b). In contrast, *ZNF646/KAT8/BCKDK* rs14235 A-carriers showed lower UPDRS-III scores compared to GG carriers (*p* < 0.05; Fig. S3c). *GCH1* rs11158026 T-carriers exhibited lower SBRs in bilateral caudate (*p* < 0.01; Fig. S3d), putamen (*p* < 0.01; Fig. S3e), and striatum (*p* < 0.01; Fig. S3f). T-carriers for both *MAPT* rs17649553 (Fig. S3g) and *LRRK2* rs76904798 (Fig. S3h) showed higher Immediate Recall scores (*p* < 0.01 and *p* < 0.05, respectively) and Derived Total Recall T-scores (*p* < 0.01 for both SNPs) of HVLT-R compared to CC carriers. *NUCKS1/Rab7L1* rs823118 T-carriers showed higher α-syn levels compared to CC carriers (*p* < 0.01; Fig. S3i). The differences in the scores of clinical assessments among genotype groups of other SNPs were not statistically different (data not shown).

### Group differences in clinical assessments among different multi-SNP configurations

Though the statistical power of associations between 14 key risk SNPs and clinical variables is low, we found several clinical variables, such as UPDRS-III scores, were associated with multiple small-effect SNPs based on multivariant regression analysis (Table 2). These findings indicate that some clinical variables of PD patients could be predicted by multiple SNPs with small effect sizes. Thus, we used ANOVA test to further examine whether patients with different genotype configurations of multiple SNPs exhibited differential clinical features. As shown in Table 2, *OGFOD2/CCDC62* rs11060180 G allele (β = 2.71, *p <* 0.01) and *GCH1* rs11158026 T allele (β = 2.27, *p <* 0.05) were associated with higher UPDRS-III scores, while *ZNF646/KAT8/BCKDK* rs14235 A allele (β = -2.30, *p <* 0.05) was associated with lower UPDRS-III scores. Thus, PD patients can be classified into 8 groups according to the genotypes of these SNPs. As shown in Figure 3a, UPDRS-III scores of PD patients were significantly different among diverse genotype configurations of *OGFOD2/CCDC62* rs11060180 (A > G: AA, AG/GG), *GCH1* rs11158026 (C > T: CC, CT/TT) and *ZNF646/KAT8/BCKDK* rs14235 (G > A: GG, GA/AA) (Fig. 3a). Those PD patients with AG/GG+CT/TT+GG genotype had highest UPDRS-III scores (28.65 ± 11.30), while PD patients with AA+CC+GA/AA genotype had lowest UPDRS-III scores (17.07 ± 8.19; Fig. 3a). The difference of UPDRS-III scores between these 2 groups was 11.58 (*p <* 0.001, One-way ANOVA test). Similarly, we found Derived Total Recall T-Scores of HVLT-R were significantly different in a gender-dependent manner among different genotype configurations of *LRRK2* rs76904798 (β = 4.07, *p <* 0.05; C > T: CC, CT/TT) and *MAPT* rs17649553 (β = 4.76, *p <* 0.01; C > T: CC, CT/TT) (Fig. 3b). Female patients with CC+CT/TT genotypes had higher Derived Total Recall T-Score compared to male patients with CC+CC genotype (*p <* 0.0001, Two-way ANOVA test). Furthermore, α-syn level in CSF were significantly different among different genotype configurations of *NUCKS1/Rab7L1* rs823118 (β = 215.40, *p <* 0.05; C > T: CC, CT/TT), *TMEM163* rs6430538 (β = -161.00, *p <* 0.05; C > T: CC, CT/TT), and *NCKIPSD* rs12497850 (β = - 188.80, *p <* 0.05; G > T: GG, GT/TT) (Fig. 3c). Patients with CT/TT+CC+GG genotype had higher α-syn level compared to patients with CC+CT/TT+GT/TT genotype who exhibited lowest α-syn level (*p <* 0.001, One-way ANOVA test).

**Figure 3.**
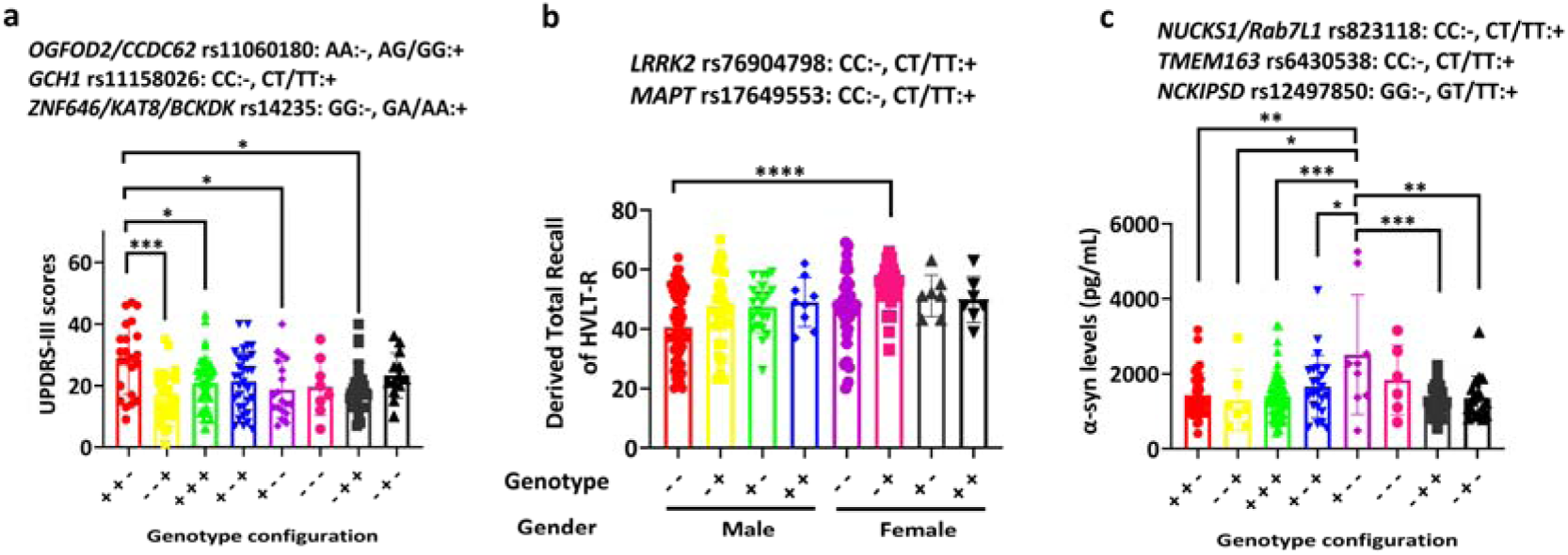
Group differences of clinical assessments among different genotype configurations. **(a)** Group difference of UPDRS-III scores among different genotype configurations (n = 23 for ++-, n = 27 for --+, n = 48 for +++, n = 32 for +-+, n = 19 for +--, n = 8 for ---, n = 25 for -++, n = 13 for -+-). **(b)** Group difference of derived total recall T-scores of HVLT-R among different genotype configurations (Male: n = 61 for --, n = 33 for -+, n = 21 for +-, n = 9 for ++; Female: n = 35 for --, n = 25 for -+, n = 7 for +-, n = 7 for ++). **(c)** Group difference of α-syn level in CSF among different genotype configurations (n = 39 for ++-, n = 7 for --+, n = 51 for +++, n = 23 for +-+, n = 9 for +--, n = 6 for ---, n = 26 for -++, n = 17 for -+-). Data were shown as mean ± SD (standard deviation). ANOVA test was used to compare the difference of clinical assessments among different genotype configurations with Bonferroni post-hoc test for multiple comparisons. *p <* 0.05 was considered statistically significant. * *p <* 0.05, ** *p <* 0.01, *** *p <* 0.001, **** *p <* 0.0001. Abbreviations, HVLT-R, Hopkins Verbal Learning Test – Revised; UPDRS-III, Unified Parkinson’s Disease Rating Scale Part III.

### Group differences of global topological metrics

As noted above, we have identified 14 key risk SNPs for PD and revealed some SNPs significantly modified the clinical assessments of PD patients. To examine whether PD-associated risk SNPs affected brain functional and structural networks, we evaluated how 14 key risk SNPs shaped the topological properties of brain networks. As shown in Figure 4, we revealed that 6 of 14 risk SNPs significantly modified the small-worldness properties of gray matter covariance network (Bonferroni-corrected *p* < 0.0036, n = 198, Two-way ANOVA test; Fig. 4a), especially for small-worldness γ and σ. The effects of 3 risk SNPs, including *OGFOD2/CCDC62* rs11060180, *GCH1* rs11158026, and *ZNF646/KAT8/BCKDK* rs14235 on global network metrics of gray matter covariance network were specifically shown in Figure 4b-d. The G-carriers (AG and GG carriers) of *OGFOD2/CCDC62* rs11060180 showed significantly lower small-worldness γ and σ of gray matter covariance network compared to AA carriers (Bonferroni-corrected *p* < 0.0036, Two-way ANOVA test; Fig. 4b). In contrast, AA carriers of *ZNF646/KAT8/BCKDK* rs14235 seemed to have higher small-worldness γ of gray matter covariance network compared to GG carriers (Bonferroni-corrected *p* < 0.0036, Two-way ANOVA test; Fig. 4d).

**Figure 4.**
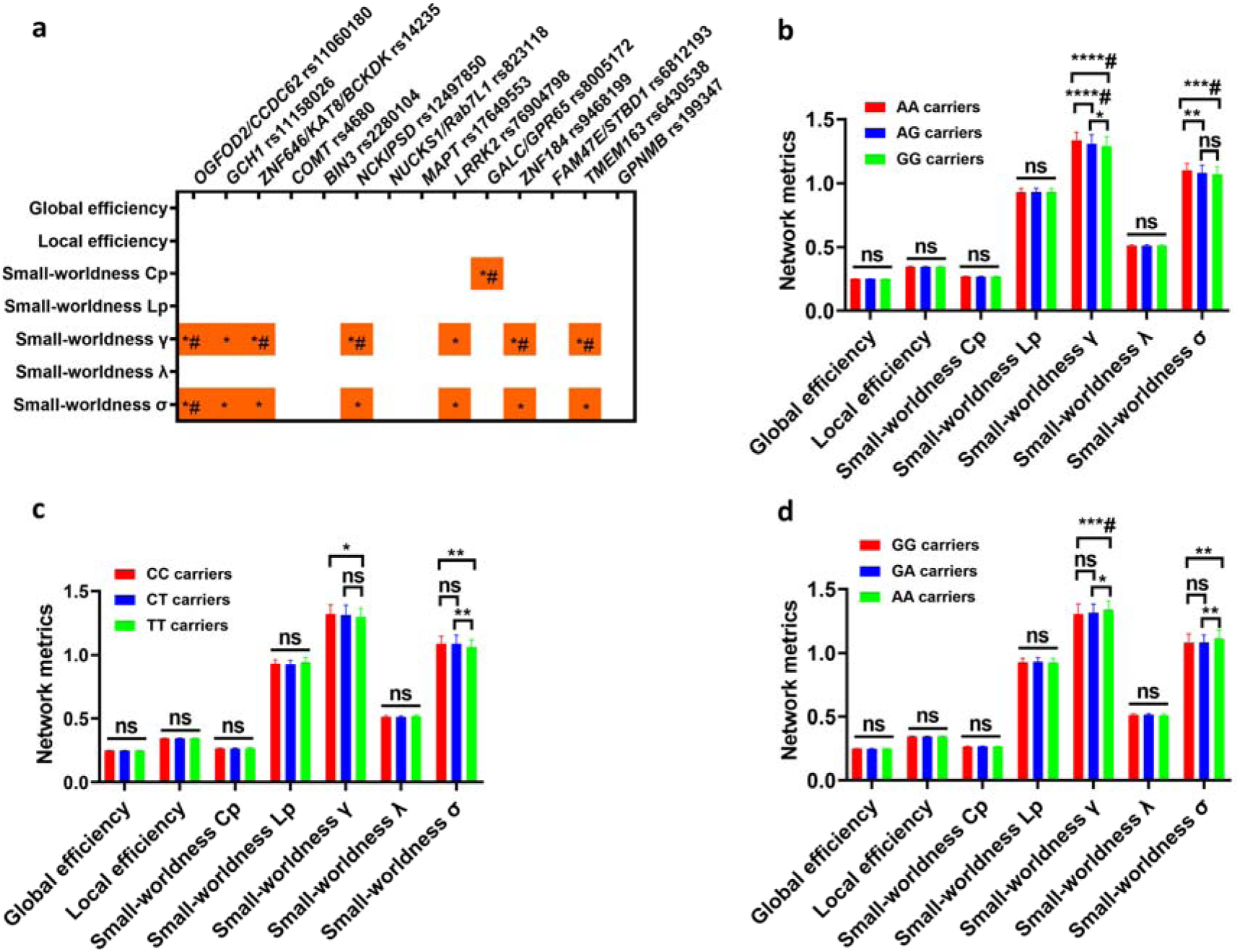
Small-worldness properties of gray matter covariance network were specifically shaped by PD-associated risk SNPs. **(a)** Group differences of global network metrics among different genotype groups (\**p* < 0.05 and ^#^*p* < 0.0036). **(b)** Group differences of global network metrics among different genotype groups of *OGFOD2/CCDC62* rs11060180. **(c)** Group differences of global network metrics among different genotype groups of *GCH1* rs11158026 (\**p* < 0.05). **(d)** Group differences of global network metrics among different genotype groups of *ZNF646/KAT8/BCKDK* rs14235 (\**p* < 0.05 and ^#^*p* < 0.0036). Two-way ANOVA test followed by Bonferroni post-hoc test was used to compare the difference of global network metrics among different genotype groups. \**p* < 0.05 and ^#^Bonferroni-corrected *p* < 0.0036 (0.05/14; 14 SNPs) were shown. Abbreviations: Cp, clustering coefficient; Lp, characteristic path length; γ, normalized clustering coefficient; λ, normalized characteristic path length; σ, small worldness.

Similarly, we found 5 of 14 risk SNPs significantly modified the small-worldness properties of white matter network (Bonferroni-corrected *p* < 0.0036, n = 146, Two-way ANOVA test; Fig. 5a). For example, AA carriers of *ZNF646/KAT8/BCKDK* rs14235 exhibited lower small-worldness γ and σ of white matter network compared to GG and GA carriers (Bonferroni-corrected *p* < 0.0036, Two-way ANOVA test; Fig. 5b). Additionally, T-carriers of *MAPT* rs17649553 showed higher small-worldness γ and σ of white matter network compared to CC carriers (Bonferroni-corrected *p* < 0.0036, Two-way ANOVA test; Fig. 5c). Furthermore, GG carriers of *GPNMB* rs199347 showed higher small-worldness γ and σ of white matter network compared to AA and AG carriers (Bonferroni-corrected *p* < 0.0036, Two-way ANOVA test; Fig. 5d).

**Figure 5.**
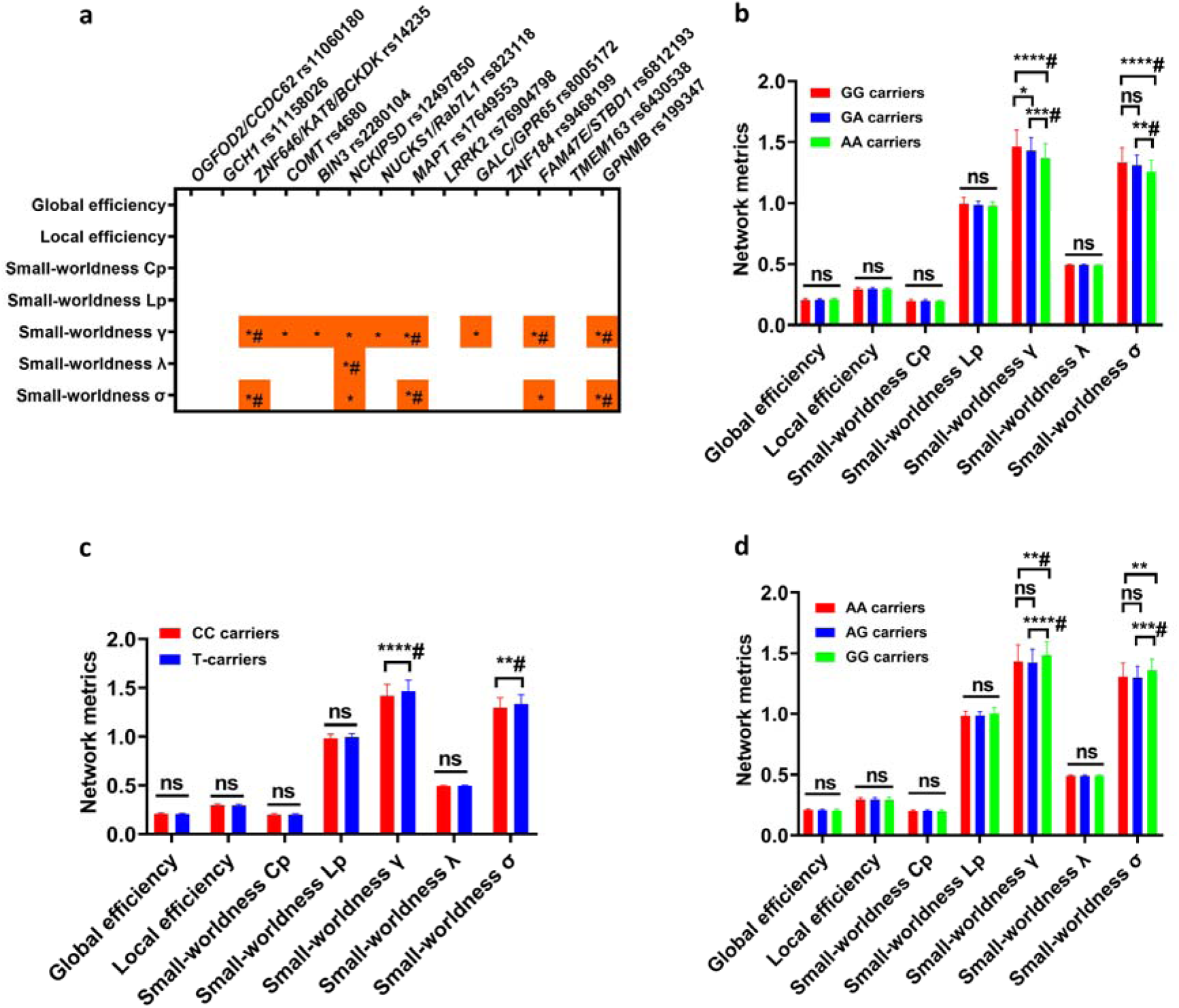
Small-worldness properties of white matter network were specifically shaped by PD-associated risk SNPs. **(a)** Group differences of global network metrics among different genotype groups (\**p* < 0.05 and ^#^*p* < 0.0036). **(b)** Group differences of global network metrics among different genotype groups of *ZNF646/KAT8/BCKDK* rs14235 (\**p* < 0.05 and ^#^*p* < 0.0036). **(c)** Group differences of global network metrics between CC carriers and T-carriers of *MAPT* rs17649553 (\**p* < 0.05 and ^#^*p* < 0.0036). **(d)** Group differences of global network metrics among different genotype groups of *GPNMB* rs199347 (\**p* < 0.05 and ^#^*p* < 0.0036). Two-way ANOVA test followed by Bonferroni post-hoc test was used to compare the difference of global network metrics among different genotype groups. \**p* < 0.05 and ^#^Bonferroni-corrected *p* < 0.0036 (0.05/14; 14 SNPs) were shown. Abbreviations: Cp, clustering coefficient; Lp, characteristic path length; γ, normalized clustering coefficient; λ, normalized characteristic path length; σ, small worldness.

For functional network, we revealed 4 of 14 risk SNPs significantly modified the small-worldness properties of functional network (Bonferroni-corrected *p* < 0.0036, n = 74, Two-way ANOVA test; Fig. S4a). For instance, AG carriers of *OGFOD2/CCDC62* rs11060180 showed higher small-worldness γ of functional network compared to AA carriers (Bonferroni-corrected *p* < 0.0036, Two-way ANOVA test; Fig. S4b). A-carriers of *ZNF646/KAT8/BCKDK* rs14235 exhibited higher small-worldness γ and σ of functional network compared to GG carriers (Bonferroni-corrected *p* < 0.0036, Two-way ANOVA test; Fig. S4c). GG carriers of *GPNMB* rs199347 showed higher small-worldness γ of functional network compared to AA carriers (Bonferroni-corrected *p* < 0.0036, Two-way ANOVA test; Fig. S4d).

### Group differences of nodal topological metrics

We also examined whether 14 risk SNPs modified the nodal metrics of brain networks. Interestingly, we found shared modifications of nodal network metrics (nodal Cp, nodal efficiency, nodal local efficiency, and nodal shortest path length) by most of 14 risk SNPs in both left caudate and right caudate of functional network, but not in white matter network and gray matter covariance network (Bonferroni-corrected *p* < 0.0008, n = 74, Two-way ANOVA test; Fig. 6a-b). For example, G-carriers of *OGFOD2/CCDC62* rs11060180 exhibited higher nodal Cp, nodal efficiency, nodal local efficiency, and lower nodal shortest path length compared to AA carriers (all Bonferroni-corrected *p* < 0.0008, Two-way ANOVA test; Fig. 6c). In addition, we found shared modifications of nodal betweenness centrality by most of 14 risk SNPs in both left putamen and right putamen of white matter network, but not in functional network and gray matter covariance network (Bonferroni-corrected *p* < 0.0036, n = 146, Two-way ANOVA test; Fig. 7a-b).

**Figure 6.**
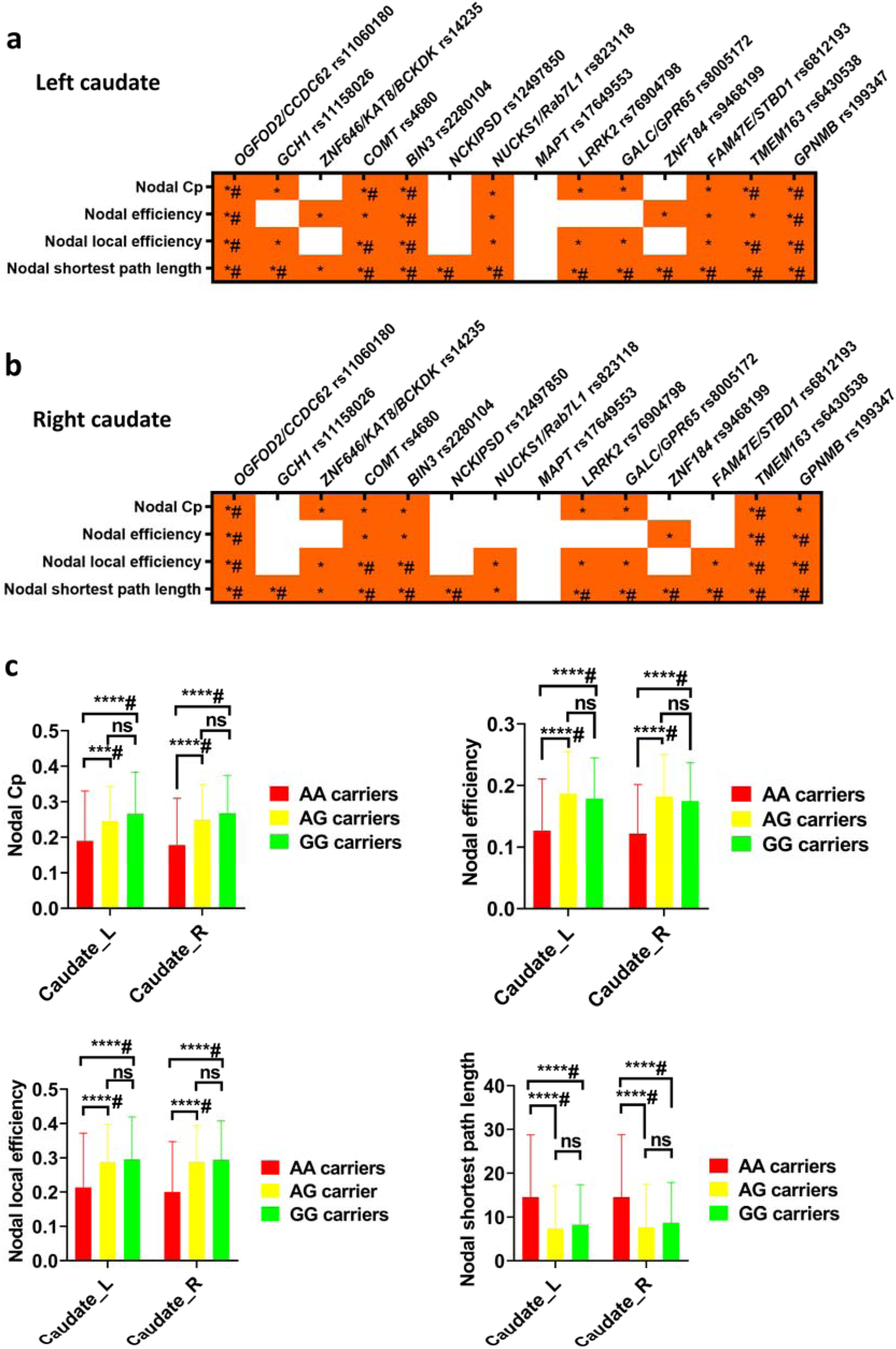
Consistent modifications of nodal metrics in bilateral caudate of functional network. **(a)** Group differences in the nodal metrics of left caudate in functional network among different genotype groups (\**p* < 0.05 and ^#^*p* < 0.0008). **(b)** Group differences in the nodal metrics of left caudate in functional network among different genotype groups (\**p* < 0.05 and ^#^*p* < 0.0008). **(c)** Group differences in the nodal metrics of bilateral caudate in functional network among different genotype groups of *OGFOD2/CCDC62* rs11060180 (\**p* < 0.05 and ^#^*p* < 0.0008). Two-way ANOVA test followed by Bonferroni post-hoc test was used to compare the difference of nodal network metrics (90 nodes) among different genotype groups. \**p* < 0.05 and ^#^Bonferroni-corrected *p* < 0.0008 (0.05/4*14; 4 nodal metrics x 14 SNPs) were shown. Abbreviations: Cp, clustering coefficient.

**Figure 7.**
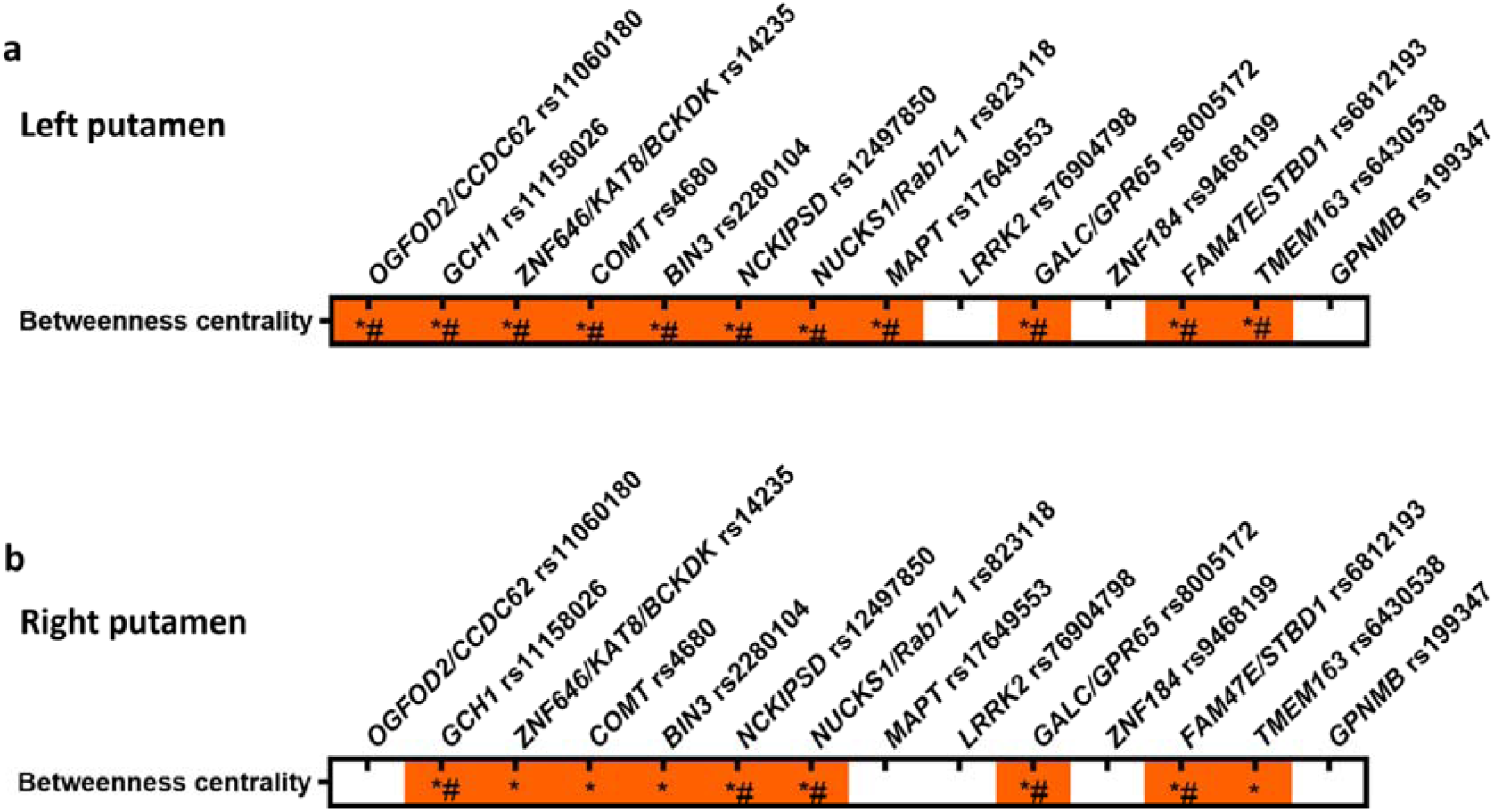
Shared modifications of nodal betweenness centrality in bilateral putamen of white matter network. **(a)** Group differences in the nodal betweenness centrality of left putamen in white matter network among different genotype groups (\**p* < 0.05 and ^#^*p* < 0.0036). **(b)** Group differences in the nodal betweenness centrality of right putamen in white matter network among different genotype groups (\**p* < 0.05 and ^#^*p* < 0.0036). Two-way ANOVA test followed by Bonferroni post-hoc test was used to compare the difference of nodal network metrics among different genotype groups. \**p* < 0.05 and ^#^Bonferroni-corrected *p* < 0.0036 (0.05/1*14; 1 nodal metrics x 14 SNPs) were shown.

### Associations between 14 risk SNPs and graphical metrics

To examine whether the effects of PD-associated risk SNPs on small-worldness properties of brain networks were independent of confounding factors, such as age, sex, disease duration, and years of education, multivariate regression analysis was performed. The Table S3 showed the associations between small-worldness properties and genotypes of risk SNPs showing statistical differences in Figure 4-5 and Figure S4. The analysis indicated that only partial risk SNPs exhibited significant associations with small-worldness properties of brain networks after Bonferroni corrections for multiple regression analysis (Table S3). We also analyzed the associations between nodal network metrics and genotypes of risk SNPs showing statistical differences in Figure 6-7, no significant associations were revealed after Bonferroni corrections for multiple regression analysis.

### Comparisons of gray matter structure and white matter integrity

We also evaluated whether 14 key risk SNPs modified the gray matter and white matter structure of PD patients. We found 14 key risk SNPs had no significant effects on the gray matter volume and cortical thickness of PD patients (Bonferroni-corrected *p >* 0.05, data not shown). Additionally, 14 key risk SNPs were also not shown to modify the integrity of white matter tracts (Bonferroni-corrected *p >* 0.05, data not shown).

### Associations between gray matter covariance network metrics and verbal memory

Because PD-associated risk SNPs significantly shaped small-worldness properties of gray matter covariance network, white matter network, and functional network, we analyzed the associations between small-worldness properties of brain network metrics and scores of clinical assessments. For small-worldness properties of gray matter covariance network, we found small-worldness γ (r = -0.21, *p* < 0.01 and β = -25.52, *p* < 0.01; Fig. 8a) and small-worldness σ (r = -0.21, *p* < 0.01 and β = -33.34, *p* < 0.01; Fig. 8b) of gray matter covariance network were negatively associated with UPDRS-III scores. In addition, small-worldness γ (r = 0.17, *p* < 0.05 and β = 4.89, *p* < 0.05; Fig. 8c) and small-worldness σ (r = 0.22, *p* < 0.01 and β = 6.65, *p* < 0.01; Fig. 8d) of gray matter covariance network were positively associated with BJLOT scores. For small-worldness properties of white matter network, we found global efficiency (r = 0.28, *p* < 0.001 and β = 63.89, *p* < 0.01; Fig. S5a), local efficiency (r = 0.32, *p* < 0.0001 and β = 34.65, *p* < 0.01; Fig. S5b), and small-worldness Cp (r = 0.26, *p* < 0.01 and β = 30.75, *p* = 0.06; Fig. S5c) of white matter network were positively associated with BJLOT scores. Additionally, small-worldness Lp (r = -0.26, *p* < 0.01 and β = -11.95, *p* < 0.01; Fig. S5d), small-worldness γ (r = -0.18, *p* < 0.05 and β = -3.27, *p* < 0.05; Fig. S5e) and small-worldness σ (r = -0.21, *p* < 0.01 and β = -4.37, *p* < 0.01; Fig. S5f) of white matter network were negatively associated with BJLOT scores.

**Figure 8.**
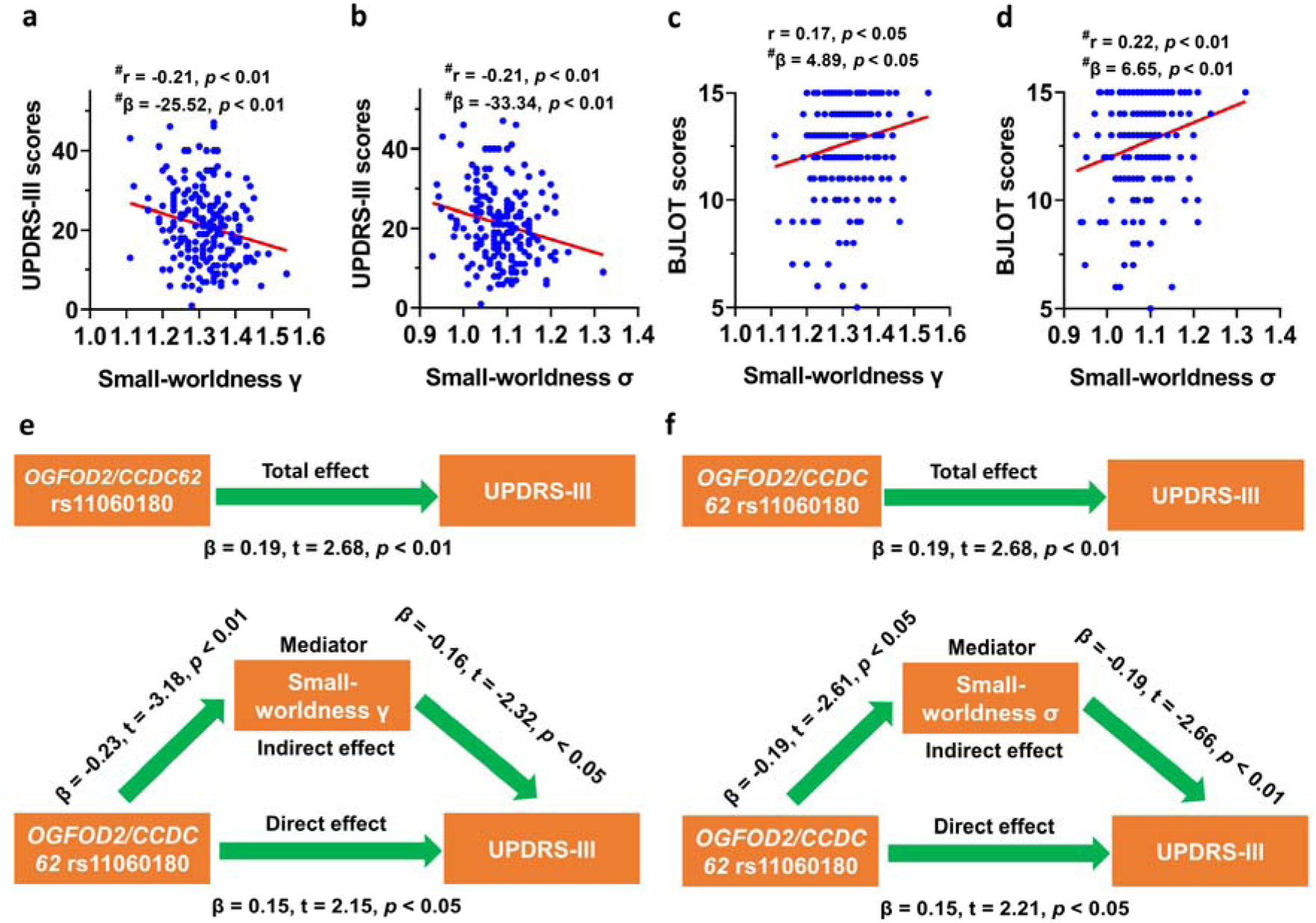
Small-worldness properties mediated the effects of *OGFOD2/CCDC62* rs11060180 on UPDRS-III scores of PD patients. **(a)** Small-worldness γ of gray matter covariance network was negatively associated with UPDRS-III scores (*p* < 0.01 in both Pearson correlation analysis and multivariate regression analysis). **(b)** Small-worldness σ of gray matter covariance network was negatively associated with UPDRS-III scores (*p* < 0.01 in both Pearson correlation analysis and multivariate regression analysis). **(c)** Small-worldness γ of gray matter covariance network was positively associated with BJLOT scores (*p* < 0.05 in both Pearson correlation analysis and multivariate regression analysis). **(d)** Small-worldness σ of gray matter covariance network was positively associated with BJLOT scores (*p* < 0.01 in both Pearson correlation analysis and multivariate regression analysis). **(e)** Mediation analysis for small-worldness γ in gray matter covariance network. **(f)** Mediation analysis for small-worldness σ in gray matter covariance network. The association analysis between graphical network metrics and UPDRS-III scores or BJLOT scores was conducted by Pearson correlation method (^#^Bonferroni-corrected *p* < 0.0125 [0.05/4]) and multivariate regression analysis with age, sex, disease duration, and years of education as covariates (^#^Bonferroni-corrected *p* < 0.0125). During the mediation analysis, age, sex, disease duration, and years of education were included as covariates. *p* < 0.05 was considered statistically significant for mediation analysis. Abbreviations: UPDRS-III, Unified Parkinson’ s Disease Rating Scale Part III; BJLOT, Benton Judgement of Line Orientation; γ: normalized clustering coefficient; σ: small worldness.

### Mediation analysis

As shown above, we revealed 3 risk SNPs, including *OGFOD2/CCDC62* rs11060180, *GCH1* rs11158026 and *ZNF646/KAT8/BCKDK* rs14235, significantly modified UPDRS-III scores (Table 2, Fig.3, Fig.S3). We also showed that small-worldness γ and small-worldness σ of gray matter covariance network were negatively associated with UPDRS-III scores (Fig. 8a-b). Thus, we examined whether small-worldness γ and small-worldness σ of gray matter covariance network mediated the effects of 3 risk SNPs (*OGFOD2/CCDC62* rs11060180, *GCH1* rs11158026 and *ZNF646/KAT8/BCKDK* rs14235) on UPDRS-III scores. Using mediation analysis, we demonstrated that AUCs of γ and σ in gray matter covariance network mediated the effects of *OGFOD2/CCDC62* rs11060180 on UPDRS-III scores (Fig. 8e-f). The mediation analysis of *GCH1* rs11158026 and *ZNF646/KAT8/BCKDK* rs14235 approached but failed to achieve a customary level of statistical significance (Fig. S6).

### Cross-validation analysis

K-fold cross-validation analysis was performed to validate the regression results shown in Fig. 8 and Figure S5. As shown in Table S4, the RMSE, R^2^, and MAE for the regression models in Figure 8 and Figure S5 were calculated by k-fold cross-validation analysis. These findings also supported that small-worldness γ and σ in gray matter covariance network were significantly associated with UPDRS-III scores and mediated the effects of *OGFOD2/CCDC62* rs11060180 on UPDRS-III scores.

## Discussion

The major finding of our study was that we revealed 14 functional SNPs conferring risk of PD in PPMI database significantly modified the topological metrics of brain functional and structural networks. Initially, we found the multi-SNP models constructed by some SNPs successfully predicted clinical assessments of PD patients. Particularly, 14 functional risk SNPs exhibited shared modifications of small-worldness properties in gray matter covariance network, white matter network, and functional network. In addition, the modifications of nodal network metrics in caudate and putamen were specifically enriched in functional network and white matter network, respectively. Furthermore, differential small-worldness properties in gray matter covariance network and white matter network modified by some SNPs were significantly correlated with UPDRS-III scores and BJLOT scores of PD patients. Finally, we demonstrated that *OGFOD2/CCDC62* rs11060180 G allele was associated with higher UPDRS-III scores and small-worldness γ and σ in gray matter covariance network mediated the effects of *OGFOD2/CCDC62* rs11060180 on UPDRS-III scores. To conclude, 14 functional risk SNPs evaluated by our study were better candidates for future functional and mechanistic studies.

To date, a large number of SNPs have been identified to be associated with the risk of PD, nevertheless, the underlying neural mechanisms of previously reported risk SNPs were largely unknown.^10,11^ Some preliminary studies have revealed PD-associated risk SNPs were associated with common pathological pathways in PD, such as endocytosis, autophagy, lysosome, mitochondria metabolism, immunological surveillance, DNA replication, synaptic vesicle recycling, and microtubule polymerization,^10,64,65^ which were consistent with our functional enrichment results (Fig. S2) based on PPI network derived from differentially expressed genes. Interestingly, we found nuclear pore complex was a novel biological pathway associated with PD-associated risk SNPs. In fact, altered nucleocytoplasmic transport has been thought as an emerging pathomechanism for multiple neurodegenerative diseases, including amyotrophic lateral sclerosis, Alzheimer disease (AD), frontotemporal dementia, and Huntington disease.^66^ It has been reported that specific nucleoporin abnormalities occurred in both sporadic and familial forms of neurodegenerative diseases and the dysfunction of nuclear pore complex contributed to disrupted nucleocytoplasmic transport.^67–69^ However, it was still unclear whether the nuclear pore complex was altered and related to the disease pathogenesis in PD, and further exploration was required.

Accumulated evidence has demonstrated that most of the traits or phenotypes in humans were affected by multiple genetic variants ^70–72^ and our study also supports this notion. Although single genetic variant in our study is not significantly associated with clinical manifestations of PD patients after stringent Bonferroni corrections (Table 2), some clinical variables, such as UPDRS-III scores and α-syn level in CSF, actually showed statistical differences among different genotype configurations of multiple SNPs (Fig. 3). Importantly, the effects of individual SNP on clinical assessments were independent of age, sex, disease duration, and genotypes of other 13 SNPs. Taken together, our novel findings support a polygenic basis underlying the clinical heterogeneity of PD patients.^11^ Because 6 SNPs (*NCKIPSD* rs12497850, *NUCKS1/Rab7L1* rs823118, *MAPT* rs17649553, *ZNF184* rs9468199, *BIN3* rs2280104, and *ZNF646/KAT8/BCKDK* rs14235) associated with clinical assessments also exhibited dramatical eQTL effects (*p <* 0.0001; Fig. S1), these SNPs deserved to be further investigated in future studies. As shown in Table 2, *OGFOD2/CCDC62* rs11060180, *GCH1* rs11158026 and *ZNF646/KAT8/BCKDK* rs14235 were significantly associated with motor symptoms of PD patients, therefore, these SNPs may have essential effects on motor symptoms of PD patients, which were demonstrated by our results showing significant differences among different genotype configurations of multi-SNP models constructed by these SNPs (Fig. 3). In fact, a previous study has shown that *GCH1* rs11158026 T-carriers had earlier age of onset by 5 years, higher UPDRS-III scores, and lower SBRs in striatum.^27^ Consistently, we also found *GCH1* rs11158026 T-carriers exhibited higher UPDRS-III scores and lower SBRs in bilateral striatum (Fig. S3 and Table 2). The association between *OGFOD2/CCDC62* rs11060180 and UPDRS-III score was also supported by a recent study showing that *OGFOD2/CCDC62* rs11060180 was associated with rapid motor progression in 365 PD patients.^73^ As shown in Table 2 and Figure S3, both *LRRK2* rs76904798 and *MAPT* rs17649553 were significantly associated with Derived Total Recall T-scores of PD patients, indicating that the verbal memory function was affected by PD-associated risk SNPs. These results were supported by our recent finding demonstrating that *MAPT* rs17649553 T allele was significantly associated with the maintenance of verbal memory in PD,^43^ in addition to its effects on PD susceptibility.^74,75^ Taken together, our results suggested that PD-associated risk SNPs indeed modified the clinical assessments of PD patients.

The disruption of network topology was an essential hallmark of PD ^76–81^ and significantly associated with motor and non-motor symptoms of PD patients.^44–48^ However, few studies have ever explored whether PD-associated risk SNPs modified the network topology of the patients. In this study, we elucidated that both global network metrics and nodal network metrics of brain networks were differentially affected by PD-associated risk SNPs. These results suggested that PD-associated risk SNP played an important role in the modifications of brain network topology. Specifically, we found small-worldness properties of both functional network and structural network were shaped by 14 risk SNPs. These results were novel and indicated that small-world topology was a shared network target of PD-associated risk SNPs. Consistently, our recent study also showed that PD-associated risk SNP, *MAPT* rs17649553, was associated with increased small-worldness γ, λ, and σ of white matter network.^43^ The small-worldness topology has been reported in a multitude of biological networks, such as gene transcriptional network,^82^ microRNA functional similarity networks,^83^ protein interaction network,^84^ and brain networks.^85^ Previous studies have shown that small-worldness topology was associated with the successful encoding of memory.^86,87^ In addition, a recent study also reported that higher small-worldness properties were associated with better working memory of patients with schizophrenia.^88^ Interestingly, the significant associations between small-worldness and memory function have been supported by other studies.^89,90^ In old adults, small-worldness properties of brain networks significantly declined compared to young people.^91^ In patients with PD, the reduction of small-worldness has been revealed,^92^ however, the potential mechanisms underlying the changes of small-worldness properties in PD remained elusive. In a recent study, we have demonstrated that age and sex significantly shaped the small-worldness properties of structural networks in PD patients,^44^ suggesting that age and sex were important individual factors modifying the small-world topology of brain networks. In current study, we provided evidence that small-world topology might also be modified by PD-associated risk SNPs, because the effects of some PD-associated risk SNPs on small-worldness properties were independent of individual demographic factors, including age, sex, disease duration, and education. Future studies were required to decipher how the PD-associated risk SNPs regulated the small-world topology of brain networks in PD.

We found *OGFOD2/CCDC62* rs11060180, *ZNF646/KAT8/BCKDK* rs14235, and *GPNMB* rs199347 exerted significant influences on global network properties of functional network and structural network, indicating that these SNPs significantly shaped both functional network and structural network. In contrast, *MAPT* rs17649553 specifically modified small-worldness properties of white matte network but not that of gray matter covariance network and functional network. These results suggested that *MAPT* rs17649553 was specifically associated with the small-world topology of white matter network, which was consistent with our recent study showing that *MAPT* rs17649553 T allele is specifically associated with higher small-world topology in white matter network of PD patients.^43^ *COMT* rs4680 seemed to be different from other SNPs due to its specific effects on the local efficiency of functional network and no effects on small-worldness properties of structural network, which were not investigated before. By contrast, three risk SNPs, including *GCH1* rs11158026, *NUCKS1/Rab7L1* rs823118, and *LRRK2* rs76904798 were not found to affect the global network metrics of functional network and structural network compared to other SNPs. To summarize, we concluded that the global network metrics were distinctly modified by 14 risk SNPs.

We elucidated that nodal network metrics of brain networks were significantly affected by 14 risk SNPs, however, the effects of 14 risk SNPs on nodal network metrics in functional network and structural network exhibited different characteristics. Specifically, the nodal metrics, such as nodal Cp, nodal efficiency, nodal local efficiency, and nodal shortest path length, in bilateral caudate of functional network were preferentially modified by 14 risk SNPs. In agreement with our results, the nodal changes of caudate nucleus in functional network of PD patients have been reported in previous studies.^80,93,94^ In contrast, the nodal betweenness centrality of bilateral putamen in white matter network was specifically shaped by PD-associated risk SNPs. In consistent with these findings, the changes of nodal network metrics in putamen of white matter network have also been revealed in PD patients.^95^ Therefore, PD-associated risk SNPs had distinct impacts on the nodal network metrics of functional network and white matter network. It should be noted that both caudate and putamen were key hubs of basal ganglia network, thus, our findings suggested that key nodes in basal ganglia network were diversely shaped by PD-associated risk SNPs. We found no significant associations between nodal network metrics and genotypes of risk SNPs using multivariate regression analysis with age, sex, disease duration, and years of education as covariates. One of the explanations for this may be that individual demographic factors, such as age and sex, have greater effects on the topological metrics of brain networks, as shown by our recent findings. ^44^

The significant correlations between graphical network metrics and clinical assessments in PD indicated that topological metrics of brain networks were predictors of clinical features in PD patients. As mentioned above, shared modifications of small-worldness properties by PD-associated risk SNPs were found in functional network and structural network, however, only small-worldness properties in structural network were significantly correlated with clinical assessments of PD patients (Fig. 8 and Fig. S5). These results indicated that small-world topological metrics in structural network but not functional network were better predictors of clinical features in PD patients. In agreement with these results, we have recently shown that small-worldness γ, λ, and σ in white matter network but not functional network were significantly correlated with verbal memory.^43^ Using mediation analysis, we demonstrated that small-worldness γ and σ in gray matter covariance network mediated the effects of *OGFOD2/CCDC62* rs11060180 on UPDRS-III scores. Therefore, graphical network metrics may provide mechanistic explanations for the effects of PD-associated risk SNPs on clinical assessments of PD patients at the network level.^43^ Although *OGFOD2/CCDC62* rs11060180, *GCH1* rs11158026 and *ZNF646/KAT8/BCKDK* rs14235 were all significantly associated with UPDRS-III scores and affected the small-worldness properties of gray matter covariance network in PD patients, only *OGFOD2/CCDC62* rs11060180 exhibited statistical significance during mediation analysis. These results suggested that small-world topology of gray matter covariance network played a more important role in the effects of *OGFOD2/CCDC62* rs11060180 on motor symptoms of PD patients. It was worth noting that small-worldness γ and σ in gray matter covariance network only partially mediated the effects of *OGFOD2/CCDC62* rs11060180 on UPDRS-III scores, which indicated that other mechanisms may also mediate the effects of *OGFOD2/CCDC62* rs11060180 on UPDRS-III scores. Future studies were required to decipher the potential molecular mechanisms underlying the effects of *OGFOD2/CCDC62* rs11060180 on motor symptoms and brain networks of PD patients.

In this study, we identified 14 PD-associated risk SNPs significantly associated with topological properties of brain networks. Among these risk genes, some of them have already been investigated before. According to previous literature, the most significant *GCH1* variant associated with PD risk is *GCH1* rs11158026 (C > T) ^96–98^ and it has been shown that rs11158026 T-carriers exhibited impaired DAT uptake and higher UPDRS-III scores.^27^ *GCH1* encoded GTP Cyclohydrolase 1(GTPCH, GCH1), which is the first and rate-limiting enzyme in the biosynthesis pathway of tetrahydrobiopterin.^99–101^ Tetrahydrobiopterin is a key cofactor required for the biosynthesis of monoamine neurotransmitters, such as serotonin, melatonin, dopamine, norepinephrine, and epinephrine.^102,103^ The dominant mutation of *GCH1* causes dopamine-responsive dystonia (DRD) or Segawa syndrome, characterized by the deficiency of catecholamines in the brain.^104–108^ Patients with DRD usually exhibited diurnally fluctuating dystonia and parkinsonian features, such as bradykinesia, tremor, rigidity, and postural instability.^104^ However, the neural mechanisms of *GCH1* variants contributing to PD risk and progression were unknown. Recently, *GCH1* deletion has been shown to reduce the expression of tyrosine hydroxylase (TH) and indirectly contribute to neuronal cell death via aberrant microglia activation.^109^ Thus, it was possible that *GCH1* polymorphisms might modify PD risk and clinical features of the patients by disrupting tetrahydrobiopterin biosynthesis, reducing dopamine availability, and exacerbating neuroinflammation in PD.^27,109^ Future studies were required to examine whether *GCH1* rs11158026 T allele was associated with impaired GCH1 enzyme activity, which was essential for its normal physiological functions.

Catechol-O-methyltransferase (COMT) is one of the key enzymes responsible for the degradation of catecholamines, including dopamine, epinephrine, and norepinephrine, as well as other substances containing a catechol structure. Levodopa, the first line therapy of PD, is also a substrate of COMT. COMT inhibitors, like entacapone and opicapone, prolong the efficacy of levodopa by inhibiting its degradation.^110,111^ *COMT* gene is enriched with multiple SNPs, such as rs6269, rs4633, rs4818, and rs4680.^112,113^ According to previous literature, rs4680 was found to be associated with executive function,^114^ obsessive-compulsive disorder,^115–117^ schizophrenia,^118,119^ anxiety,^120,121^ addiction,^122,123^ bipolar disorder,^124,125^ suicide,^126,127^ depression,^128,129^ psychosis in AD,^130^ and treatment response of levodopa therapy in PD.^131,132^ Therefore, the changes of COMT activity due to single nucleotide variation may affect brain function and disease susceptibility. In PD, *COMT* rs4680 was associated with fronto-cortical dopamine turnover,^133^ striatal denervation,^134^ gray matter atrophy,^135^ age at onset,^136^ executive function,^131,137–140^ attention control,^141^ treatment response of COMT inhibitors,^23^ levodopa response variability,^142^ dyskinesias,^143^ pain,^144^ wearing-off phenomenon,^145,146^ and mild cognitive impairment.^24^ In our study, we found *COMT* rs4680 was associated with LNS scores (β = 0.65, *p* = 0.0119) and affected global and nodal metrics of functional network. Future studies were required to understand how *COMT* rs4680 modified the clinical manifestations and brain network metrics of PD patients.

*GPNMB* rs199347 has been demonstrated to be a key genetic locus of PD and associated with α-syn pathology ^10,12^ and the expression of GPNMB is selectively enhanced in the substantia nigra of PD patients and increases after lysosomal stress.^147^ According to GTEx database, *GPNMB* rs199347 G allele was significantly associated with reduced expression of *GPNMB* in multiple brain regions, including cortex (*p <* 1.8 x 10^-20^), caudate (*p <* 3.0 x 10^-19^), putamen (*p <* 5.5 x 10^-24^), and substantia nigra (*p <* 2.1 x 10^-5^). In addition, *GPNMB* rs199347 G allele was also significantly associated with reduction of *NUPL2* expression in cortex (*p <* 5.0 x 10^-17^), caudate (*p <* 6.6 x 10^-10^), putamen (*p <* 5.6 x 10^-8^), and substantia nigra (*p <* 1.3 x 10^-11^). Consistently, Diaz-Ortiz et al. (2022) demonstrated that individuals carrying *GPNMB* rs199347 haplotype exhibited allele-specific expression for *GPNMB*.^12^ They also confirmed that moderate to high levels of GPNMB protein are expressed in the human brain of multiple postmortem cases by immunoblotting and immunohistochemistry.^12^ Because higher expression of GPNMB conferred pathogenicity in PD,^12^ G allele was protective for the reductions of *GPNMB* levels in G-carriers of PD. Considering that GPNMB expression was found in multiple cell types of the brain,^12^ future studies were required to dissect its role in the modulation of brain function.

We revealed that *NCKIPSD* rs12497850 specifically affected the small-worldness properties of gray matter covariance network and white matter network. How this genetic variant modified structural networks has not been explored before. *NCKIPSD* encodes SPIN90 protein, which is highly expressed in synapses.^148–150^ Recent studies have shown that SPIN90 protein is involved in the endocytosis of synaptic vesicles and also critical for the formation of normal dendritic spines.^148–151^ Phosphorylated SPIN90 enhances neuronal synaptic activity by interacting with PSD95 and Shank proteins.^151^ Whereas activation of NMDA receptor induces the dephosphorylation of SPIN90 and initiates cofilin-mediated actin assembly and dendritic spine contraction.^152^ Kim *et al*. (2017) reported that SPIN90 regulates long-term depression and hippocampus-dependent behavioral flexibility.^153^ A recent study further revealed that SPIN90 regulates the assembly of cortical actin by interacting with mDia1 and Arp2/3 complexes.^154^ Taken together, it was possible that *NCKIPSD* rs12497850 might modify structural networks in PD patients by regulating the development of cerebral cortex and the morphology of neurite processes.

It remained unknown about the molecular mechanisms underlying the effects of *MAPT* rs17649553 T allele on verbal memory and small-wordlness properties of white matter network. According to the eQTL data, *MAPT* rs17649553 was associated with differential expressions of multiple genes, including *ARL17A*, *CRHR1*, *KANSL1*, *LRRC37A*, and *MAPT*. Interestingly, these genes have been reported to be associated with multiple neuropsychiatric diseases.^155–160^ In addition, *MAPT* rs17649553 was also associated with the splicing of *CRHR1*, *KANSL1*, *MAPT*, *PLEKHM1*, and *ARHGAP27*. The detailed associations between *MAPT* locus and PD have been discussed in our recent study.^43^ Future studies were required to decipher the molecular mechanisms underlying the effects of *MAPT* rs17649553 on the risk, small-world topology, and verbal memory of PD patients. *LRRK2* is an essential risk gene for PD and G2019S was the most common genetic mutation of *LRRK2* that was widely investigated in previous decades.^161–164^ In this study, we found *LRRK2* rs76904798 was significantly associated with verbal memory. This finding was novel and deserved to be further validated with a larger sample size of PD patients. The associations between *GALC/GPR65* rs8005172 and clinical features or brain networks of PD patients have been not investigated so far. A recent study reported that another *GALC* variant rs979812 affected the enzymatic activity of galactosylceramidase and risk of PD patients.^165^ It seemed that it was increased but not reduced galactosylceramidase activity that causally associated with PD.^165^ Additionally, GALC was hypothesized to modulate the clearance of misfolded α-syn aggregates through autophagic-lysosomal pathway.^166^ Whether altering the activity of galactosylceramidase could be utilized as a therapeutic target of PD should be further explored.

Compared to above risk SNPs, *OGFOD2/CCDC62* rs11060180, *BIN3* rs2280104, *NUCKS1/Rab7L1* rs823118, *ZNF184* rs9468199, *FAM47E/STBD1* rs6812193, *TMEM163* rs6430538, and *ZNF646/KAT8/BCKDK* rs14235 were less investigated in previous literature. Future studies were required to decode the neural mechanisms underlying the effects of these SNPs on the clinical manifestations and network metrics of PD patients.

In current study, we revealed motor and non-motor symptoms could be successfully predicted by multi-SNP models constructing by some PD-associated risk SNPs, suggesting that these risk SNPs may have potential clinical utility to predict and monitor the clinical manifestations of PD patients. In addition, this study systematically evaluated how multiple PD-associated risk SNPs affected brain network metrics. The relevant findings may provide potential explanations for the significant associations between these SNPs and PD risk at the network level. The specific effects of PD-associated risk SNPs on the small-world topology of gray matter covariance network, white matter network, and functional network indicate that PD-associated risk SNPs preferentially shaped the global small-world properties of brain networks. Particularly, the enrichment of nodal changes in bilateral caudate and putamen of basal ganglia network suggested that key hubs in basal ganglia network were essential targets for PD-associated risk variants. Furthermore, our mediation analysis provided potential network mechanisms to explain the effects of *OGFOD2/CCDC62* rs11060180 on UPDRS-III scores. Taken together, the results revealed by our study deepened our understanding of the potential associations between PD-associated risk SNPs and clinical characteristics and brain networks of PD patients. The limitation of this study was that we didn’t investigate the molecular mechanisms underlying the effects of PD-associated risk SNPs on clinical features and brain networks of PD patients. Future studies were needed to explore how these risk SNPs affect the pathophysiology of PD at the molecular level and whether these risk genes could be utilized to develop potential diagnostic biomarkers and therapeutic targets for PD.

To summarize, we found 14 PD-associated SNPs, including *OGFOD2/CCDC62* rs11060180, were eQTLs and associated with multiple biological pathways involved in endocytosis-autophagy-lysosome, mitochondria, gene transcription, and nuclear pore complex. Interestingly, PD patients with specific combinations of some SNPs exhibited statistically different clinical features. In addition, we revealed both shared and distinct brain network metrics were significantly shaped by PD-associated genetic variants. Small-worldness properties at the global level and nodal metrics in caudate and putamen of basal ganglia network were preferentially modified. Finally, we demonstrated that small-worldness properties in gray matter covariance network mediated the effects of *OGFOD2/CCDC62* rs11060180 on motor severity of PD patients.

## Data Availability

All the raw data used in the preparation of this Article were downloaded from PPMI database (www.ppmi-info.org/data). All data produced in the present study are available upon reasonable request to the authors.

## Contributors

All authors read and approved the final version of the manuscript. Zhichun Chen: Conceptualization, Formal analysis, Visualization, Methodology, Writing - original draft, Writing - review and editing; Bin Wu: Validation, Investigation, Methodology; Guanglu Li: Data curation, Formal analysis, Visualization; Liche Zhou: Data curation, Formal analysis, Investigation; Lina Zhang: Formal analysis, Investigation, Methodology; Jun Liu: Conceptualization, Supervision, Funding acquisition, Writing - original draft, Project administration, Writing- review and editing.

## Data sharing statement

The data supporting the findings of this study are available from the corresponding author upon reasonable request. All clinical, biochemical, genetic, and imaging data are deposited in PPMI database and can be accessed at www.ppmiinfo.org.

## Acknowledgements

This work was supported by grants from the National Key Research and Development Program (2016YFC1306505), the National Natural Science Foundation of China (81471287, 81071024, 81171202). These funding sources had no role in study design, conduct, analysis, interpretation or writing of the manuscript, or in the decision to submit the manuscript. We thank the share of PPMI data by all the PPMI study investigators. For up-to-date information on PPMI study, visit www.ppmiinfo.org. PPMI – a public-private partnership – is funded by the Michael J. Fox Foundation for Parkinson’s Research and funding partners, which can be found at www.ppmiinfo.org/fundingpartners.

## Supplementary materials

Supplementary data related to this article can be found online.

---

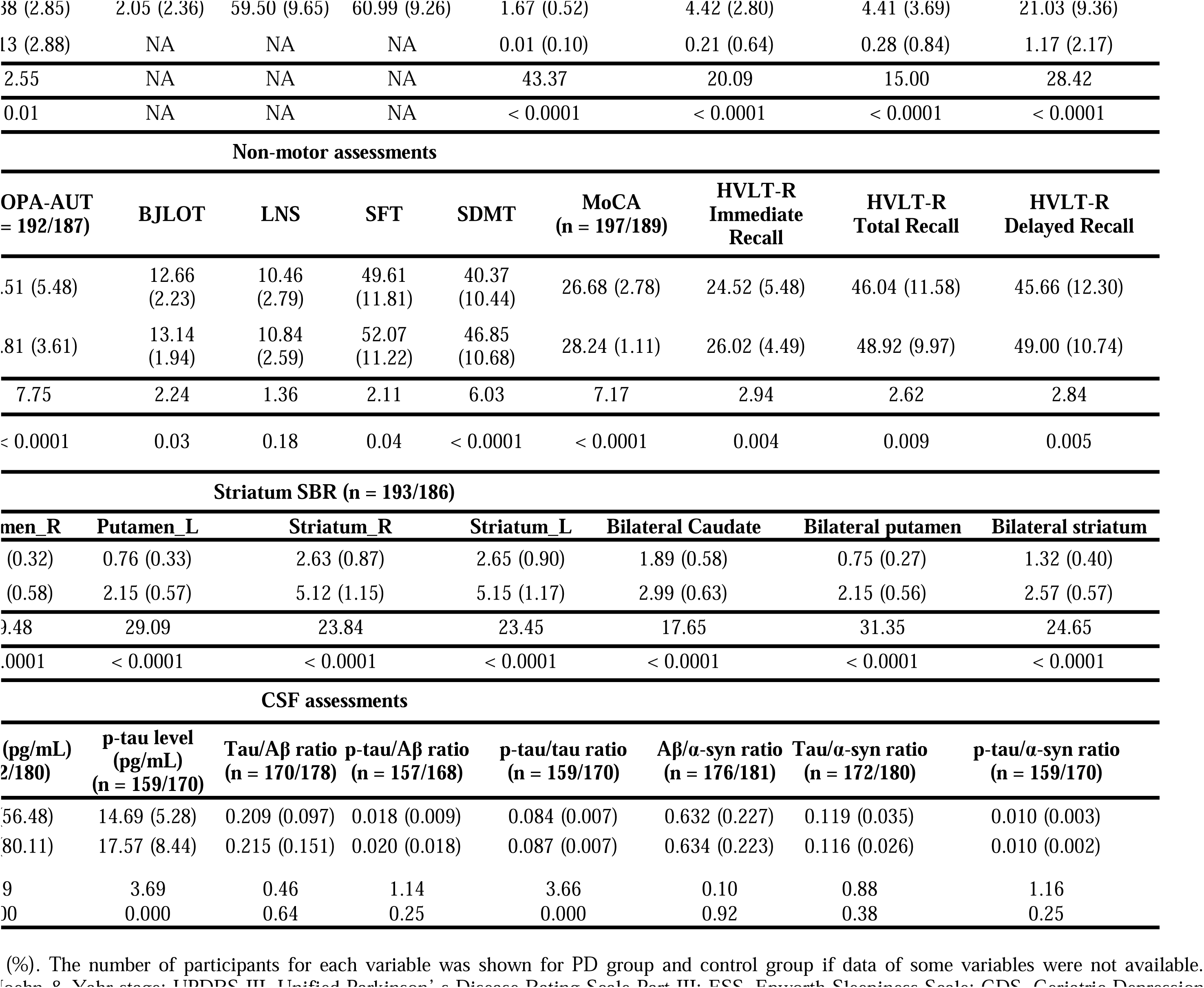
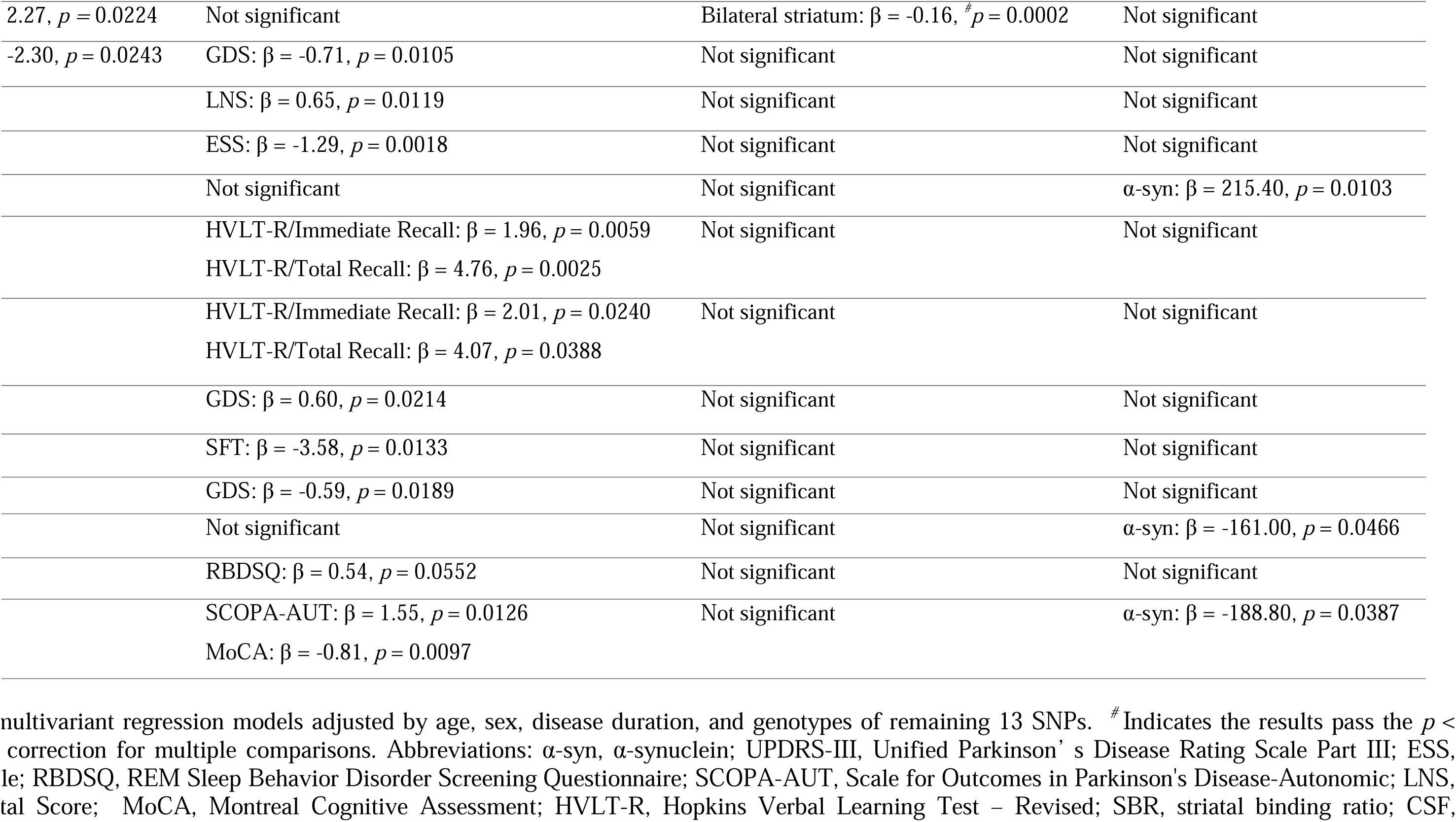

