## Supplemental file for "Parkinson’s disease-associated genetic variants synergistically shape brain networks"

**Supplementary Material**

**Table S1-S4**

**Figure S1-S6**

| SNP | Location | OR （(95%CI) | *p* value | Reference |
| --- | --- | --- | --- | --- |
| *OGFOD2/CCDC62* rs11060180 | chr12:123303586 | 0.90 (0.88–0.92) | 2.05 x 10^-20^ | Nature genetics (2017) ^1^ |
| *GCH1* rs11158026 | chr14:55348869 | 0.91 (0.89–0.93) | 4.30 x 10^-16^ | Nature genetics (2017) ^1^ |
| *ZNF646/KAT8/BCKDK* rs14235 | chr16:31121793 | 1.08 (1.06–1.10) | 5.44 x 10^−12^ | Nature genetics (2017) ^1^ |
| *COMT* rs4680 | chr22: 19963748 | 0.60 (0.41–0.90) | 1.50 x 10^-2^ | Pharmacogenetics and Genomics (2012) ^2^ |
| *BIN3* rs2280104 | chr8:22525980 | 1.07 (1.04–1.09) | 2.53 x 10^−8^ | Nature genetics (2017) ^1^ |
| *NUCKS1* rs823118 | chr1:205723572 | 0.89 (0.87–0.91) | 1.12 x 10^−23^ | Nature genetics (2017) ^1^ |
| *MAPT* rs17649553 | chr17:43994648 | 0.78 (0.76–0.80) | 1.26 x 10^−68^ | Nature genetics (2017) ^1^ |
| *LRRK2* rs76904798 | chr12:40614434 | 1.15 (1.12–1.19) | 1.21 x 10^−19^ | Nature genetics (2017) ^1^ |
| *GALC/**GPR65* rs8005172 | chr14:88472612 | 1.08 (1.05–1.10) | 8.77 x 10^−11^ | Nature genetics (2017)^1^ |
| *ZNF184* rs9468199 | chr6:27681215 | 1.11 (1.08–1.14) | 1.46 x 10^−12^ | Nature genetics (2017) ^1^ |
| *FAM47E/STBD1* rs6812193 | chr4:77198986 | 0.92 (0.90–0.94) | 1.43 x 10^−14^ | Nature genetics (2017) ^1^ |
| *TMEM163* rs6430538 | chr2:135539967 | 0.89 (0.87–0.91) | 8.24 x 10^−24^ | Nature genetics (2017) ^1^ |
| *GPNMB* rs199347 | chr7:23293746 | 0.91 (0.89–0.93) | 3.51 x 10^−18^ | Nature genetics (2017) ^1^ |
| *NCKIPSD/CCDC71/IP6K2* rs12497850 | chr3:48748989 | 0.93 (0.91–0.96) | 9.16 x 10^−9^ | Nature genetics (2017) ^1^ |

***Table S1*: The genome location, OR, *p* values, and reference of 14 SNPs.**

Abbreviations: OR, odds ratio; SNP, single nucleotide polymorphism; *OGFOD2*, 2-Oxoglutarate And Iron Dependent Oxygenase Domain Containing 2; *GCH1*, GTP Cyclohydrolase 1; *ZNF646,* Zinc Finger Protein 646; *KAT8,* Lysine Acetyltransferase 8; *BCKDK,* Branched Chain Keto Acid Dehydrogenase Kinase; *COMT,* Catechol-O-Methyltransferase; *BIN3,* Bridging Integrator 3; *NUCKS1,* Nuclear Casein Kinase And Cyclin Dependent Kinase Substrate 1; *MAPT,* Microtubule Associated Protein Tau; *LRRK2,* Leucine Rich Repeat Kinase 2; *GALC,* Galactosylceramidase; *GPR65,* G Protein-Coupled Receptor 65; *ZNF184, Zinc Finger Protein 184; FAM47E,* Family With Sequence Similarity 47 Member E; *STBD1,* Starch Binding Domain 1; *TMEM163*, transmembrane protein 163; *GPNMB*, Glycoprotein Nmb; *NCKIPSD*, NCK Interacting Protein With SH3 Domain; *CCDC71,* Coiled-Coil Domain Containing 71; *IP6K2*, Inositol Hexakisphosphate Kinase 2.

***Table S2*: The demographic and clinical data for each group of subjects.**

| Clinical variable | Group 1 | Group 2 | Group 3 | *p* - value |
| --- | --- | --- | --- | --- |
| Age, years | 61.55 ± 9.26 | 61.24 ± 9.21 | 62.34 ± 8.21 | *p* > 0.05 |
| Sex (Male/Female) | 124/74 | 93/53 | 47/27 | *p* > 0.05 |
| Education, years | 15.38 ± 2.85 | 15.27 ± 2.99 | 15.09 ± 2.82 | *p* > 0.05 |
| Disease duration, years | 2.05 ± 2.36 | 2.04 ± 2.15 | 1.96 ± 2.45 | *p* > 0.05 |
| HY | 1.64 ± 0.52 | 1.66 ± 0.50 | 1.68 ± 0.53 | *p* > 0.05 |
| Tremor | 4.04 ± 3.48 | 3.82 ± 3.26 | 4.06 ± 3.67 | *p* > 0.05 |
| Rigidity | 4.24 ± 2.81 | 4.29 ± 2.82 | 4.28 ± 2.75 | *p* > 0.05 |
| UPDRS-III | 21.03 ± 9.36 | 20.85 ± 9.64 | 20.45 ± 10.30 | *p* > 0.05 |
| RBDSQ | 4.17 ± 2.68 | 4.25 ± 2.67 | 4.37 ± 2.75 | *p* > 0.05 |
| SCOPA-AUT | 9.51 ± 5.48 | 9.94 ± 5.63 | 10.86 ± 5.58 | *p* > 0.05 |
| LNS | 10.46 ± 2.79 | 10.55 ± 2.87 | 10.11 ± 2.81 | *p* > 0.05 |
| BJLOT | 12.66 ± 2.23 | 12.74 ± 2.16 | 12.65 ± 2.02 | *p* > 0.05 |
| SFT | 49.61 ± 11.81 | 49.32 ± 11.38 | 49.95 ± 11.93 | *p* > 0.05 |
| SDMT | 40.37 ± 10.44 | 40.45 ± 10.90 | 38.73 ± 11.22 | *p* > 0.05 |
| MoCA | 26.68 ± 2.78 | 26.98 ± 2.69 | 26.56 ± 2.90 | *p* > 0.05 |
| Derived Total Recall of HVLT-R | 46.04 ± 11.58 | 45.40 ± 12.08 | 45.49 ± 13.85 | *p* > 0.05 |
| Derived Delayed Recall of HVLT-R | 45.66 ± 12.30 | 45.35 ± 12.51 | 45.68 ± 13.79 | *p* > 0.05 |
| SBRs of Caudate_R | 1.88 ± 0.61 | 1.80 ± 0.54 | 1.74 ± 0.59 | *p* > 0.05 |
| SBRs of Caudate_L | 1.89 ± 0.63 | 1.78 ± 0.59 | 1.73 ± 0.63 | *p* > 0.05 |
| SBRs of Putamen_R | 0.75 ± 0.32 | 0.74 ± 0.32 | 0.72 ± 0.37 | *p* > 0.05 |
| SBRs of Putamen_L | 0.76 ± 0.33 | 0.73 ± 0.33 | 0.70 ± 0.35 | *p* > 0.05 |
| SBRs of Striatum_R | 2.46 ± 0.90 | 2.54 ± 0.81 | 2.46 ± 0.90 | *p* > 0.05 |
| SBRs of Striatum_L | 2.65 ± 0.90 | 2.51 ± 0.87 | 2.44 ± 0.92 | *p* > 0.05 |
| SBRs of Bilateral Caudate | 1.89 ± 0.58 | 1.79 ± 0.53 | 1.74 ± 0.57 | *p* > 0.05 |
| SBRs of Bilateral putamen | 0.75 ± 0.27 | 0.74 ± 0.28 | 0.71 ± 0.32 | *p* > 0.05 |
| SBRs of Bilateral striatum | 1.32 ± 0.40 | 1.26 ± 0.38 | 1.23 ± 0.42 | *p* > 0.05 |
| Aβ level (pg/mL) | 865.4 ± 375.9 | 830.3 ± 330.3 | 809.2±342.9 | *p* > 0.05 |
| α-syn level (pg/mL) | 1491.0 ± 752.2 | 1438.0 ± 715.5 | 1429.0 ± 636.1 | *p* > 0.05 |
| Tau level (pg/mL) | 166.5 ± 56.48 | 163.7 ± 53.15 | 166.6 ± 57.89 | *p* > 0.05 |
| p-tau level (pg/mL) | 14.69 ± 5.28 | 14.32 ± 4.84 | 14.65 ± 5.28 | *p* > 0.05 |

The data were shown as the mean ± standard deviation (SD). The motor function examination was assessed in ON state. The comparisons of clinical variables among three groups were performed with one-way ANOVA test followed by Bonferroni post-hoc test. Abbreviations: Aβ, β-amyloid; α-syn, α-synuclein; HY, Hoehn & Yahr stage; UPDRS-III, Unified Parkinson’ s Disease Rating Scale Part III; RBDSQ, REM Sleep Behavior Disorder Screening Questionnaire; SCOPA-AUT, Scale for Outcomes in Parkinson's Disease-Autonomic; SDMT, Symbol Digit Modalities Test; LNS, Letter Number Sequencing; SFT, Semantic Fluency Test Score; BJLOT, Benton Judgement of Line Orientation; MoCA, Montreal Cognitive Assessment; HVLT-R, Hopkins Verbal Learning Test – Revised; SBR, striatal binding ratio; CSF, cerebrospinal fluid.

***Table S3*: The associations between 14 risk SNPs and small-worldness properties.**

| Gray matter covariance network | | | | |
| --- | --- | --- | --- | --- |
| Metrics  SNPs | **Small-worldness γ** | **Small-worldness σ** | | |
| *OGFOD2/CCDC62* rs11060180 | **β = -0.0235, ^#^*p* = 0.0017** | β = -0.0160, *p* = 0.0098 | | |
| *ZNF646/KAT8/BCKDK* rs14235 | β = 0.0146, *p* = 0.0686 | - | | |
| *NCKIPSD* rs12497850 | β = 0.0019, *p* = 0.8199 | - | | |
| *FAM47E/STBD1* rs6812193 | β = 0.0021, *p* = 0.7772 | - | | |
| *TMEM163* rs6430538 | β = 0.0053, *p* = 0.4545 | - | | |
| Statistical significance: ^#^Bonferroni-corrected *p* < 0.0083 (0.05/6 test) | | | | |
| White matter network | | | | |
| Metrics  SNPs | **Small-worldness γ** | **Small-worldness λ** | | **Small-worldness σ** |
| *ZNF646/KAT8/BCKDK* rs14235 | **β = -0.0438, ^#^*p* = 0.0057** | **-** | | **β = -0.0366, ^#^*p* = 0.0049** |
| *NCKIPSD* rs12497850 | **-** | **β = 0.3451, ^#^*p* < 0.0001** | | **-** |
| *MAPT* rs17649553 | β = 0.0430, *p* = 0.0294 | - | | β = 0.0320, *p* = 0.0485 |
| *FAM47E/STBD1* rs6812193 | β = 0.0015, *p* = 0.9150 |  | | **-** |
| *GPNMB* rs199347 | β = 0.0194, *p* = 0.1974 |  | | β = 0.0167, *p* = 0.1787 |
| Statistical significance: ^#^Bonferroni-corrected *p* < 0.0063 (0.05/8 test) | | | | |
| Functional network | | | | |
| Metrics  SNPs | **Small-worldness γ** | | **Small-worldness σ** | |
| *OGFOD2/CCDC62* rs11060180 | β = 0.0348, *p* = 0.1696 | | β = 0.0211, *p* = 0.3272 | |
| *ZNF646/KAT8/BCKDK* rs14235 | **β = 0.0774, ^#^*p* = 0.0048** | | **β = 0.0664, ^#^*p* = 0.0043** | |
| *GPNMB* rs199347 | β = 0.0090, *p* = 0.2505 | | - | |
| Statistical significance: ^#^Bonferroni-corrected *p* < 0.01 (0.05/5 test) | | | | |

The data are shown as the β and *p*-values derived from multivariant regression models adjusted by age, sex, disease duration, and years of education. *^#^* Indicates *p*-values pass Bonferroni corrections for multiple statistical tests. Abbreviations: γ, normalized clustering coefficient; λ, normalized characteristic path length; σ, small worldness, SNP, single nucleotide polymorphism.

***Table S4*: K-fold cross-validation analysis.**

| Cross-validation analysis for the results shown in Figure 8 | | | |
| --- | --- | --- | --- |
| Validation metrics | **RMSE** | **R^2^** | **MAE** |
| Small-worldness γ (UPDRS-III correlation) | 9.0446 | 0.1014 | 7.3189 |
| Small-worldness σ (UPDRS-III correlation) | 9.0040 | 0.1060 | 7.3288 |
| Small-worldness γ (BJLOT correlation) | 1.9336 | 0.2714 | 1.5368 |
| Small-worldness σ (BJLOT correlation) | 1.9277 | 0.2858 | 1.5278 |
| Small-worldness γ (Mediation analysis) | 8.9802 | 0.1000 | 7.2634 |
| Small-worldness σ (Mediation analysis) | 8.9281 | 0.1107 | 7.2151 |
| Cross-validation analysis for the results shown in Figure S5 | | | |
| Validation metrics | **RMSE** | **R^2^** | **MAE** |
| Global efficiency | 1.9044 | 0.2347 | 1.5381 |
| Local efficiency | 1.9115 | 0.2377 | 1.5119 |
| Small-worldness Cp | 1.9371 | 0.2338 | 1.5247 |
| Small-worldness Lp | 1.9135 | 0.2312 | 1.5433 |
| Small-worldness γ | 1.9234 | 0.2230 | 1.5515 |
| Small-worldness σ | 1.9145 | 0.2300 | 1.5472 |

Abbreviations: UPDRS-III, Unified Parkinson’ s Disease Rating Scale Part III; BJLOT, Benton Judgement of Line Orientation; RMSE, Root mean squared error; R², coefficient of determination; MAE, mean absolute error; Cp, clustering coefficient; Lp, characteristic path length; γ, normalized clustering coefficient; σ, small worldness.

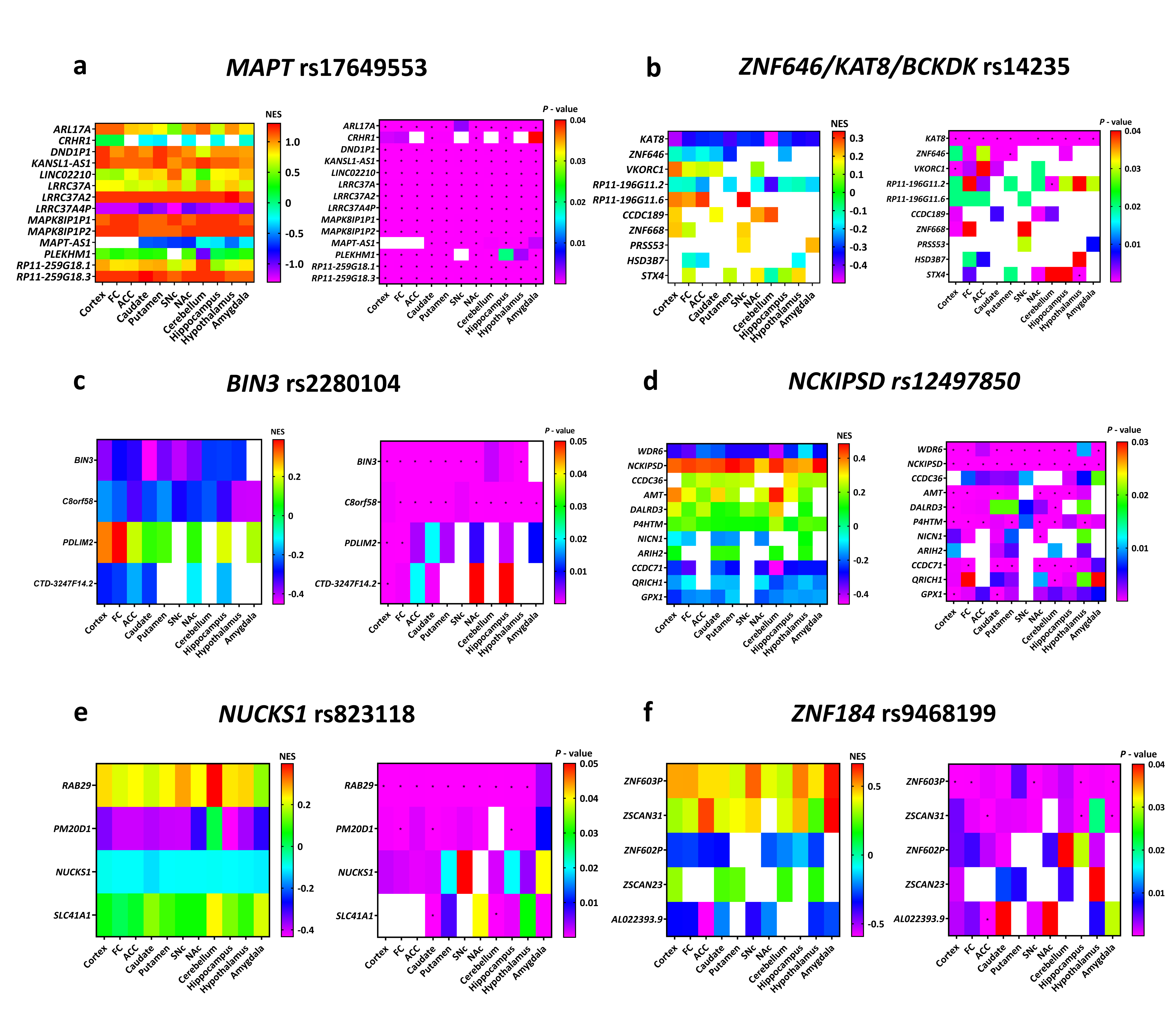

**Figure S1.** The eQTL effects of 6 representative SNPs associated with PD. **(a)-(f)** NES and *p-*values for altered gene expressions associated with *MAPT* rs17649553 **(a)**, *ZNF646/KAT8/BCKDK* rs14235 **(b)**, *BIN3* rs2280104 **(c)**, *NCKIPSD* rs12497850 **(d)**, *NUCKS1/Rab7L1* rs823118 **(e)**, and *ZNF184* rs9468199 **(f)** in eQTL analysis (**p* < 0.0001). Abbreviations: ACC, anterior cingulate cortex; *AMT*, aminomethyltransferase; *ARHGAP27,* Rho GTPase activating protein 27; *ARIH2*, ariadne RBR E3 ubiquitin protein ligase 2; *ARL17A*, ADP ribosylation factor like GTPase 17A; *BIN3*, bridging integrator 3; *CCDC36*, coiled-coil domain containing 36; *CCDC71*, coiled-coil domain containing 71; *CCDC189*, coiled-coil domain containing 189; *CCNT2*, cyclin T2; *C8orf58*, chromosome 8 open reading frame 58; *CRHR 1*, corticotropin releasing hormone receptor 1; *DALRD3*, DALR anticodon binding domain containing 3; *DND1P1*, DND microRNA-mediated repression inhibitor 1 pseudogene 1; eQTL, expression quantitative trait loci; FC, frontal cortex; *GPX1* = glutathione peroxidase 1; *HSD3B7*, hydroxy-delta-5-steroid Dehydrogenase, 3 beta- and steroid delta-isomerase 7; *IP6K2*, inositol hexakisphosphate kinase 2; *KANSL1*, lysine acetyltransferase 8 regulatory NSL complex subunit 1; *KANSL1-AS1*, lysine acetyltransferase 8 regulatory NSL complex subunit 1 antisense RNA 1; *KAT8*, lysine acetyltransferase 8; *LRRC37A*, leucine rich repeat containing 37A; *LRRC37A2*, leucine rich repeat containing 37 member A2; *MAPT*, microtubule-associated protein tau; *MAPT-AS1*, microtubule-associated protein tau antisense RNA 1; *LINC02210*, long intergenic non-protein coding RNA 2210; *MAPK8IP1P1*, mitogen-activated protein kinase 8 interacting protein 1 pseudogene 1; *MAPK8IP1P2*, mitogen-activated protein kinase 8 interacting protein 1 pseudogene 2; NAc, nucleus accumbens; *NCKIPSD*, NCK interacting protein with SH3 domain; NES, normalized effect size; *NICN1*, Nicolin 1; *NUCKS1*, nuclear casein kinase and cyclin dependent kinase substrate 1; *P4HTM*, prolyl 4-hydroxylase, transmembrane; *PM20D1*,peptidase M20 domain containing 1; *PRSS53*, serine protease 53; *PLEKHM1*, pleckstrin homology and RUN domain containing M1; *PDLIM2*, PDZ and LIM domain 2; *QRICH1*, glutamine rich 1; *RP11-259G18.1*, retinitis pigmentosa-11-259G18.1; *RP11-259G18.3*, retinitis pigmentosa-11-259G18.3; *RP11-196G11.2*, retinitis pigmentosa-11-196G11.2; *RP11-196G11.6*, retinitis pigmentosa-11-196G11.6; *LRRC37A4P*, leucine rich repeat containing 37 member A4 pseudogene; *RAB29*, Rab GTPase protein 29; *SLC41A1*, solute carrier family 41 member 1; SNc, substantia nigra; *STX4*, syntaxin 4; *VKORC1*, vitamin K epoxide reductase complex subunit 1; *WDR6*, WD repeat domain 6; *ZNF603P*, zinc finger protein 603 pseudogene; *ZNF602P*, zinc finger protein 602 pseudogene; *ZNF646*, zinc finger protein 646; *ZNF668*, zinc finger protein 668; *ZSCAN23*, zinc finger and SCAN domain containing 23; *ZSCAN31*, zinc finger and SCAN domain containing 31.

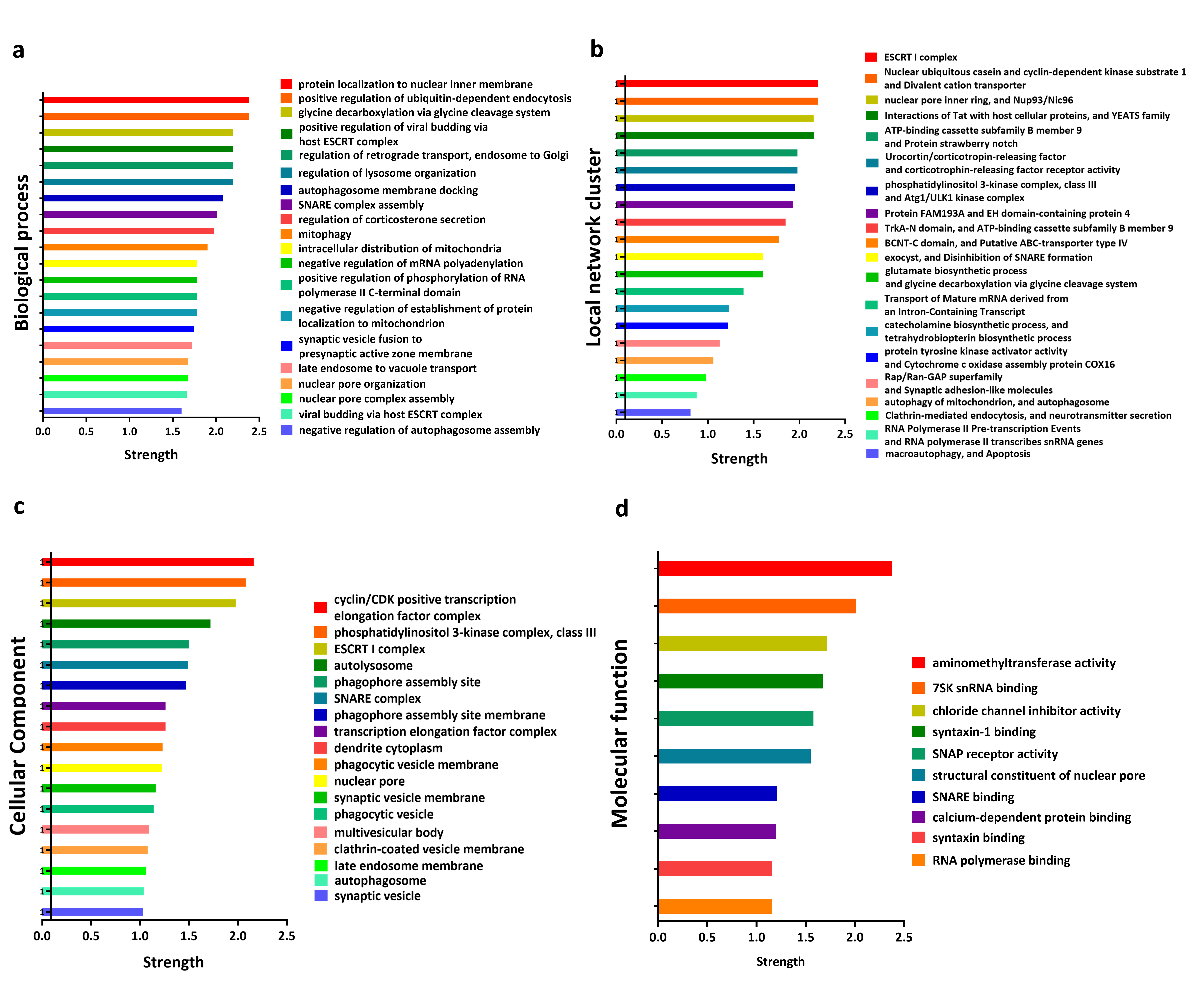
 **Figure S2.** Functional enrichment analysis of differentially expressed genes associated with 14 SNPs. **(a)-(d)** Functional enrichment analysis was performed to reveal the biological processes **(a)**, local network cluster **(b)**, cellular component **(c)**, and molecular function **(d)** of differentially expressed genes associated with 14 SNPs based on STRING database (*p* < 0.05, FDR corrected).

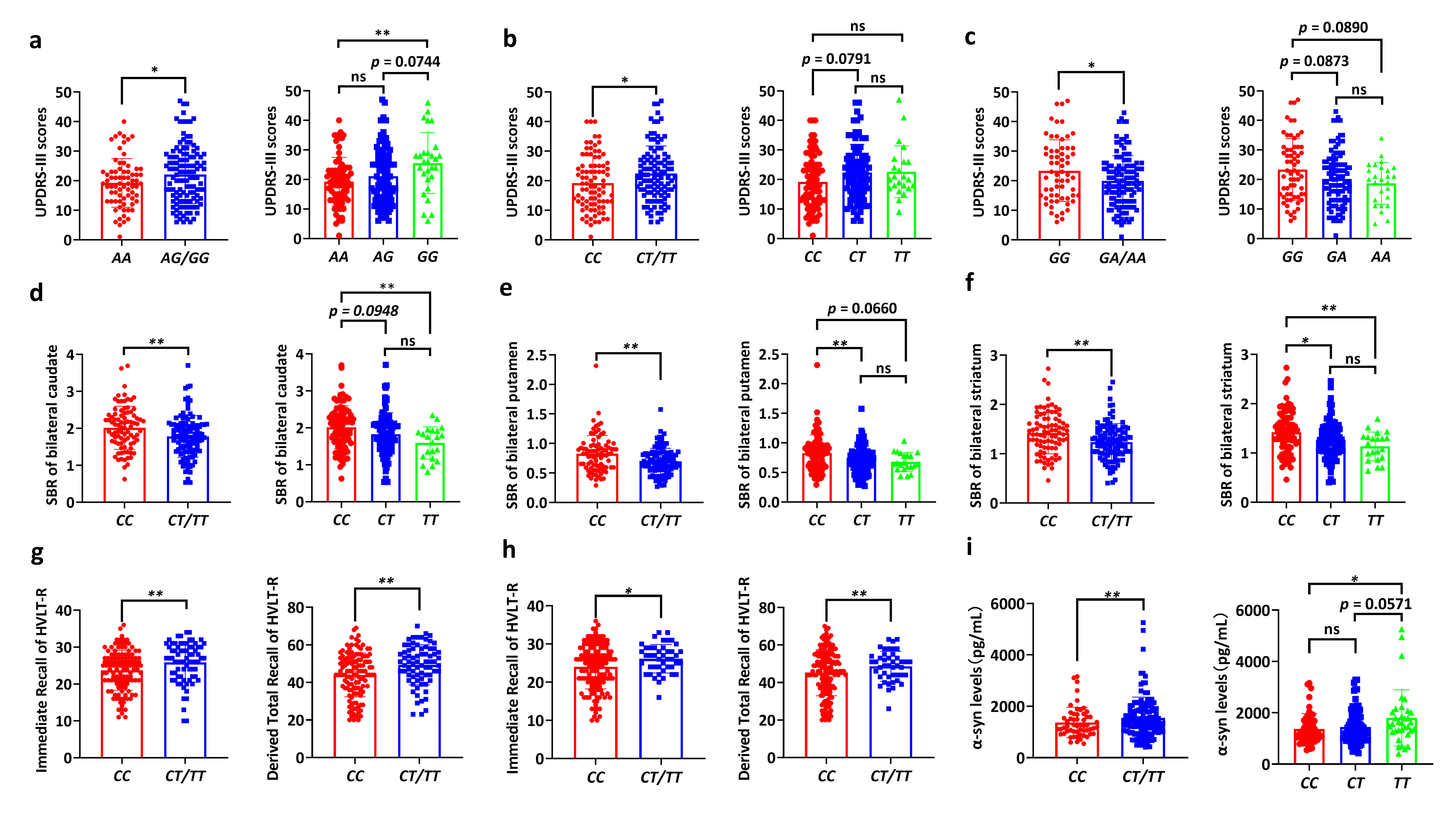
**Figure S3.** Group differences of clinical assessments among different SNP genotypes. Group differences of UPDRS-III scores among different genotypes of *OGFOD2/CCDC62* rs11060180 **(a)**, *GCH1* rs11158026 **(b)**, and *ZNF646/KAT8/BCKDK* rs14235 **(c)**. Group difference of SBRs in bilateral caudate **(d)**, putamen **(e)**, and striatum **(f)** among different genotypes of *GCH1* rs11158026. Group differences of Immediate Recall scores and Derived Total Recall T-scores for different genotypes of *MAPT* rs17649553 **(g)** and *LRRK2* rs76904798 **(h)**. Group differences of α-syn levels for different genotypes of *NUCKS1/Rab7L1* rs823118 **(i)**. Unpaired t-test (for two groups comparison) and one-way ANOVA test followed by Bonferroni post-hoc test (for three groups comparison) were used to compare the difference of clinical assessments among different SNP genotypes. *p <* 0.05 was considered statistically significant. * *p <* 0.05, ** *p <* 0.01. Abbreviations: UPDRS-III, Unified Parkinson’ s Disease Rating Scale Part III; HVLT-R, Hopkins Verbal Learning Test – Revised; SBR, striatal binding ratio; α-syn, α-synuclein.

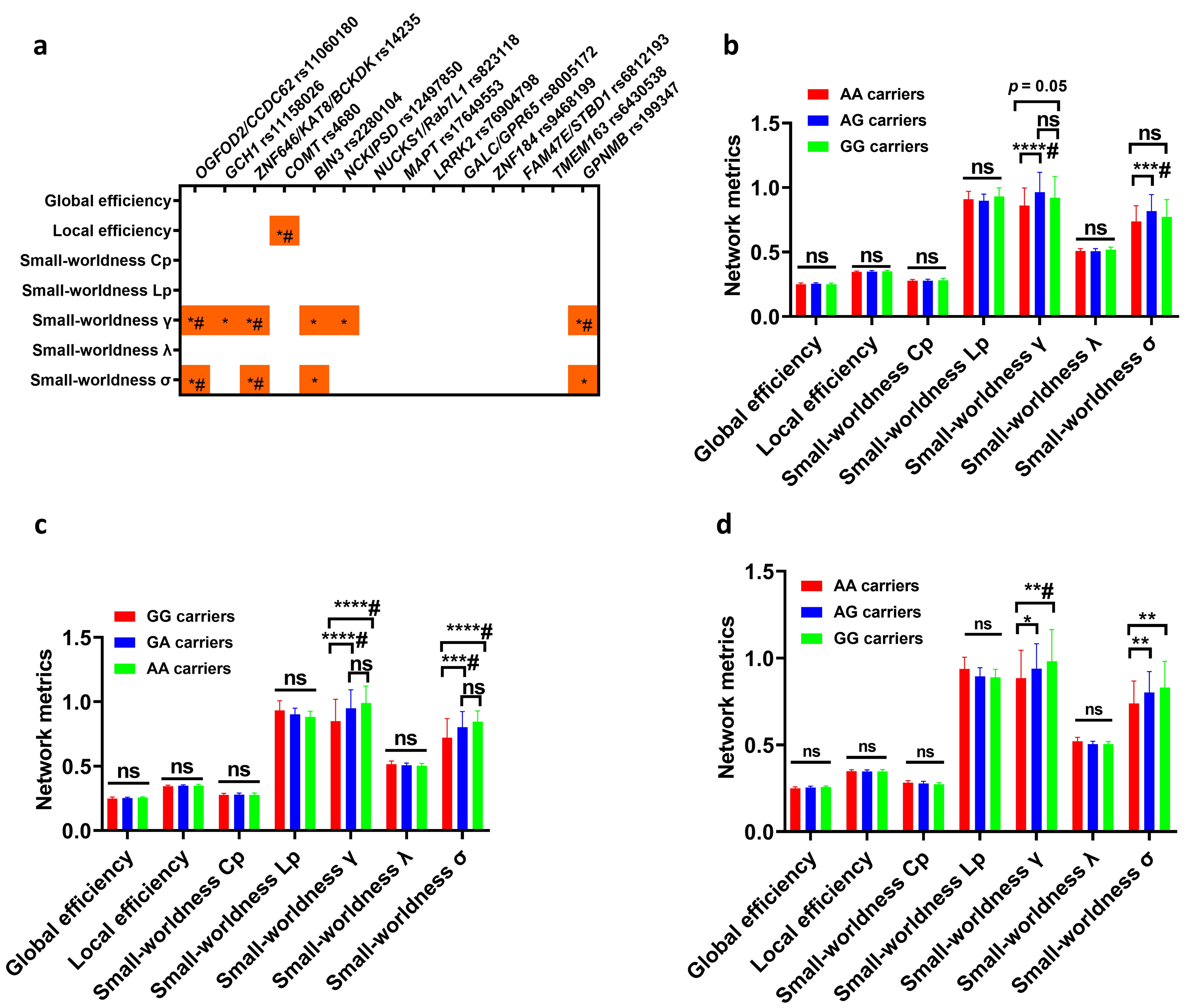
 **Figure S4. Group differences in the global network metrics of functional network.** **(a)** Group differences of global network metrics among different genotype groups (**p* < 0.05 and ^#^*p* < 0.0036). **(b)** Group differences of global network metrics among different genotype groups of *OGFOD2/CCDC62* rs11060180 (**p* < 0.05 and ^#^*p* < 0.0036). **(c)** Group differences of global network metrics among different genotype groups of *ZNF646/KAT8/BCKDK* rs14235 (**p* < 0.05 and ^#^*p* < 0.0036). **(d)** Group differences of global network metrics among different genotype groups of *GPNMB* rs199347 (**p* < 0.05 and ^#^*p* < 0.0036). Two-way ANOVA test followed by Bonferroni post-hoc test was used to compare the difference of global network metrics among different genotype groups. **p* < 0.05 and ^#^Bonferroni-corrected *p* < 0.0036 (0.05/14; 14 SNPs) were shown. Abbreviations: Cp: clustering coefficient; Lp: characteristic path length; γ: normalized clustering coefficient; λ: normalized characteristic path length; σ: small worldness.

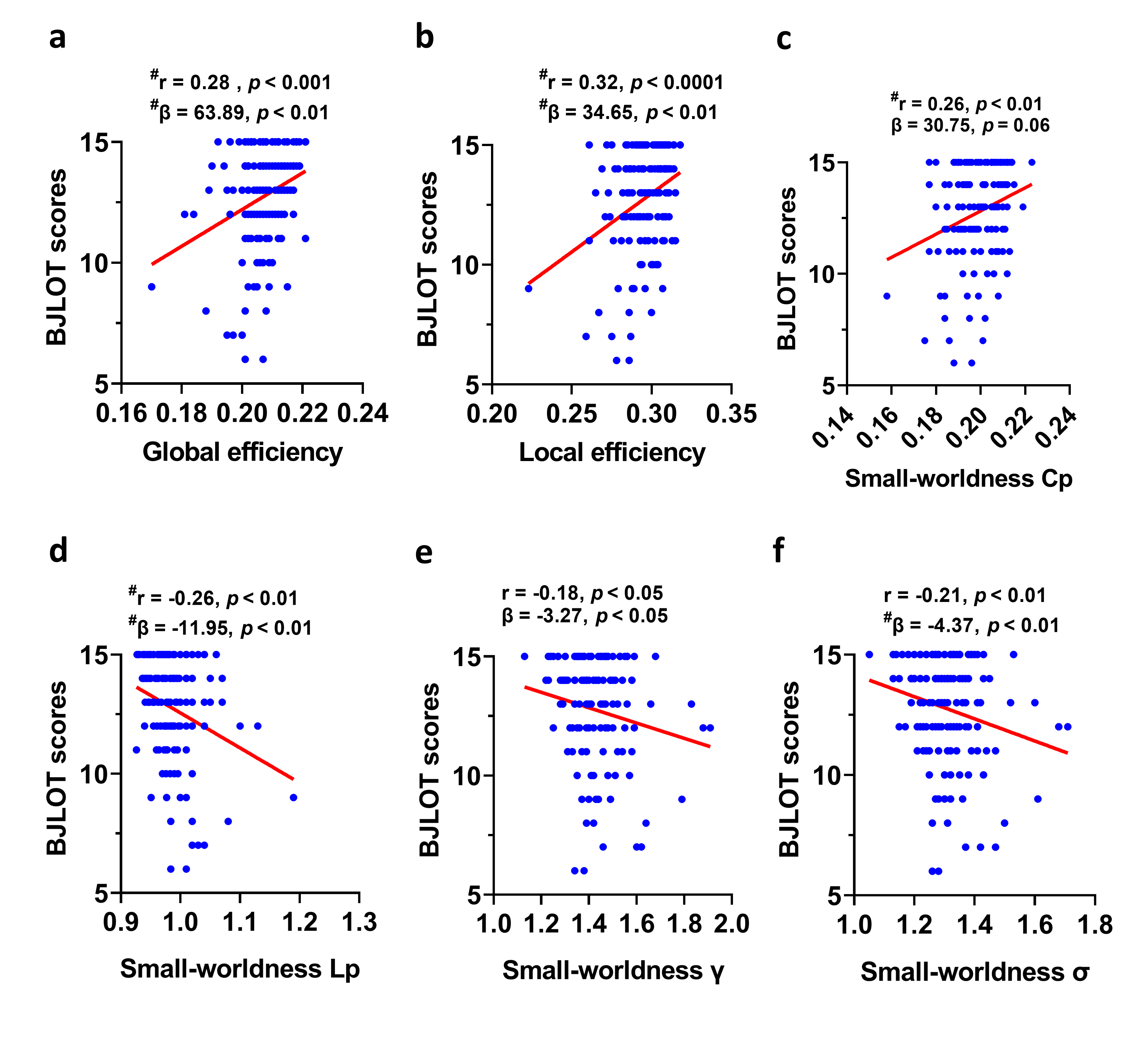
**Figure S5. The associations between global network metrics of white matter network and BJLOT scores. (a)** Global efficiency of white matter network was positively associated with BJLOT scores (*p* < 0.001 in Pearson correlation analysis and *p* < 0.01 in multivariate regression analysis). **(b)** Local efficiency of white matter network was positively associated with BJLOT scores (*p* < 0.0001 in Pearson correlation analysis and *p* < 0.01 in multivariate regression analysis). **(c)** Small-worldness Cp of white matter network was positively associated with BJLOT scores (*p* < 0.01 in Pearson correlation analysis and *p* = 0.06 in multivariate regression analysis). **(d)** Small-worldness Lp of white matter network was negatively associated with BJLOT scores (*p* < 0.01 in Pearson correlation analysis and *p* < 0.01 in multivariate regression analysis). **(e)** Small-worldness γ of white matter network was negatively associated with BJLOT scores (*p* < 0.05 in Pearson correlation analysis and *p* < 0.05 in multivariate regression analysis). **(f)** Small-worldness σ of white matter network was negatively associated with BJLOT scores (*p* < 0.01 in Pearson correlation analysis and *p* < 0.01 in multivariate regression analysis). The association analysis between graphical network metrics and BJLOT scores was conducted by Pearson correlation method (^#^Bonferroni-corrected *p* < 0.0083 [0.05/6]) and multivariate regression analysis with age, sex, disease duration, and years of education as covariates (^#^Bonferroni-corrected *p* < 0.0083). Results with uncorrected *p <* 0.05 and Bonferroni-corrected results were reported. Abbreviations: BJLOT, Benton Judgement of Line Orientation; γ: normalized clustering coefficient; λ: normalized characteristic path length; σ: small worldness.

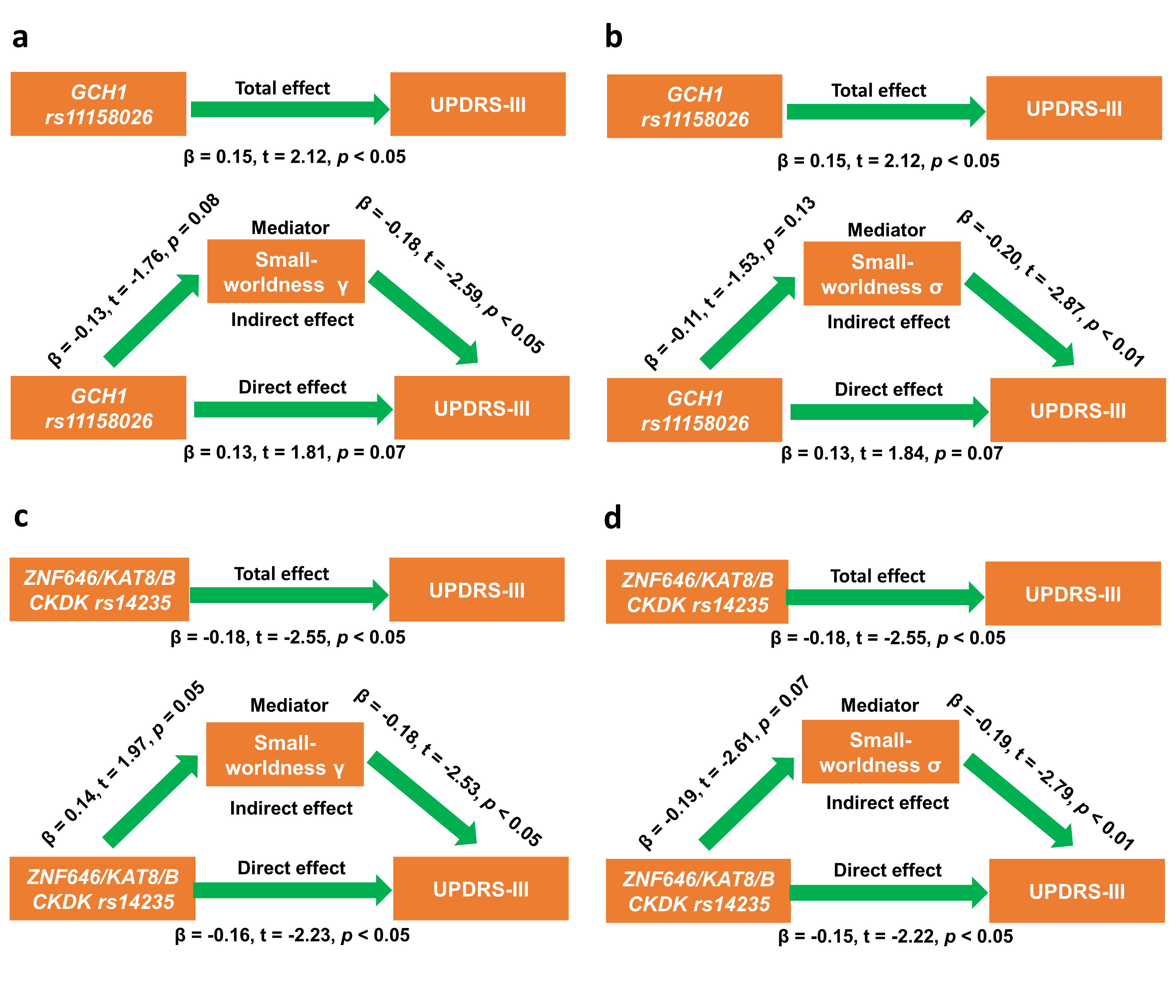
 **Figure S6.** Mediation analysis for *GCH1* rs11158026 and *ZNF646/KAT8/BCKDK* rs14235. **(a)** Small-worldness γ didn’t mediate the effects of *GCH1* rs11158026 on UPDRS-III scores. **(b)** Small-worldness σ didn’t mediate the effects of *GCH1* rs11158026 on UPDRS-III scores. **(c)** Small-worldness γ didn’t mediate the effects of *ZNF646/KAT8/BCKDK* rs14235 on UPDRS-III scores. **(d)** Small-worldness σ didn’t mediate the effects of *ZNF646/KAT8/BCKDK* rs14235 on UPDRS-III scores. During the mediation analysis, age, sex, disease duration, and years of education were included as covariates. *p* < 0.05 was considered statistically significant. Abbreviations: UPDRS-III, Unified Parkinson’ s Disease Rating Scale Part III; AUC: Area under curve; γ: normalized clustering coefficient; σ: small worldness.
